# Reporting quality in psychological interventions. Implications for cumulative science and evidence-based practice: A scoping review

**DOI:** 10.64898/2025.11.26.25341096

**Authors:** J. Vives, S. Lorente, A. Casellas, E. López-González, J.A. Marín-García, C. Molla, M.F. Rodrigo, P. Viguer, J.-M. Losilla

## Abstract

**Background:** Poor quality in research reporting is a persistent issue that has hindered the advancement of scientific knowledge across all health science disciplines. Similarly, the progress of evidence-based practice has been hampered, as the replication of interventions is often impeded by the incomplete reporting of implementation details.

**Objective:** To map how reports of psychological interventions describe implementation at both the whole intervention level and the level of recipients’ behaviours or actions (BeA), and to examine whether reporting quality varies by field, guideline use, or indicators of intervention complexity.

**Methods:** This scoping review followed PRISMA-ScR. Searches were conducted on PsycINFO (ProQuest) and Web of Science (Clarivate) for publications (2014-2024) describing the development and/or implementation of psychological interventions in clinical, public health, social/organizational or educational fields. A random sample of 200 publications per field was screened for inclusion. Following a two-stage selection process involving double screening and consensus resolution, 80 studies were included. Data were extracted using a Delphi-based instrument and analysed using descriptive statistics and χ² tests to compare different aspects of reporting across fields, guideline use, and complexity features.

**Results:** Only 11.3% reported development-only objectives, while most combined development with effectiveness (40%) and feasibility (48.8%). Reporting quality tools were seldom used (13.8%). Interventions typically targeted a single recipient category (87.5%); multilevel targeting was rare (6.3%). Implementation sequence was often replicable (73.8%), but structured aids were underused (tables/lists 38.8%; diagrams 16.3%). Costs were rarely reported (3.8%). At the BeA level, structural features were usually replicable (e.g., type/form of administration, periodicity, place, timing), whereas content/materials (36.3% and 47.5%), adaptation/customization (35%), and fidelity/adherence (27.5%) were frequently incomplete. Few significant between field differences emerged. Reporting tended to improve as the number of BeA increased.

**Conclusions:** Psychological intervention reports commonly omit critical replicability details, especially provider/evaluator prerequisites, content/materials, adaptation rules, fidelity, and costs, despite available guidance. Routine use of structured representations (e.g., flow diagrams, BeA tables) and established reporting tools (e.g., TIDieR, CONSORT-SPI) could close the most consequential gaps and facilitate replication and implementation at scale.

**Registration:** INPLASY 2025.7.0098.

## Introduction

Evidence-based practice is grounded in cumulative science, which is usually based on systematic reviews of the results of primary studies. However, the ability of a systematic review to synthesize evidence is often reduced, or even precluded, by deficiencies in the reporting of primary studies (Turner et al., 2012; Losilla et al., 2018). Poor quality in research reporting, whether due to a lack of completeness, transparency and/or precision, is a pressing problem that has hindered the advancement of scientific knowledge across all health science disciplines for decades (Glasziou et al., 2014; Rosengaard et al., 2024). Moreover, inadequate reporting of research not only may prevent its replication, but it is also a major source of research waste, which in areas such as biomedicine is estimated to account for 85% of research resources (Ioannidis et al., 2014; Glasziou & Chalmers, 2018; Almaqrami et al., 2020; Blanco et al., 2020).

The quality of reporting in primary studies focusing on an intervention, whether concerning its development, feasibility, efficacy, effectiveness or efficiency, is not expected to differ. In the case of interventions, however, another key implication arises in relation to evidence-based practice. If the characteristics related to the implementation of proposed psychological interventions are not reported with sufficient information, it is not possible to faithfully replicate them. Consequently, any practice involving the application of such interventions cannot truly be considered evidence based.

The emergence of reporting guidelines, such as CONSORT (Begg, 1996; Moher, Schulz & Altman, 2001; Schulz et al., 2010; Hopewell et al., 2025), STROBE (Vandenbroucke et al., 2007), PRISMA (Liberati et al., 2009; Page et al., 2021), SPIRIT (Chan et al., 2013, 2025) or TREND (Des Jarlais, Lyles & Crepaz, 2004), has led to an improvement in the quality of research reporting. Nevertheless, the impact of these guidelines and authors’ adherence to them remain in need of improvement (Bastuji-Garin et al., 2013; Mannocci et al., 2014; Clayson et al., 2019; Orduña-Malea et al., 2023). In the case of interventions, the recent publication of guidelines to develop interventions, such as those focusing on complex interventions of Medical Research Council (MRC) (Craig et al., 2008; Tanner-Smith & Grant, 2018; Skivington et al., 2021) and the Agency for Healthcare Research and Quality (AHRQ) (Guise et al., 2017b), and the availability of intervention-specific reporting tools and guides such as CReDECI 2 (Möhler, Köpke & Meyer, 2015; Tanner-Smith & Grant, 2018), GUIDED (Duncan et al., 2020), TIDieR (Hoffmann et al., 2014), iCAT_SR (Lewin et al., 2017) and RIPI-f (Lopez-Alcalde et al., 2022), may have contributed to some improvement in the reporting quality of interventions.

Note that some of these guides target complex interventions and address how the factors contributing to their complexity should be reported to enable accurate replication and evaluation. Taking the factors that define the complexity of an intervention into account allows for a more thorough description of the characteristics of published interventions and to explore whether these factors, which can be challenging to report appropriately, are related to the reporting quality. Although several definitions of complex interventions attempt to distinguish them from non-complex ones, complexity is neither a binary characteristic nor a property of a single type of intervention (Petticrew, 2011; Lewin et al., 2017). Rather, any intervention can be placed on a continuum ranging from a low degree to a high degree of complexity (Moore et al., 2017). While the debate surrounding the concept of ‘complex intervention’ is still ongoing, the most influential conceptualizations published in recent years, such as those of the MRC and the AHRQ aforementioned, agree that, in an intervention, complexity may arise, first and foremost, from the properties of the intervention itself, being the presence of multiple components, and the relations between them and the outcomes (especially moderations and mediations) the most relevant properties, along with the adaptability or customizability of the protocol.

Partitioning the intervention into components may facilitate the study of its efficacy and effectiveness (Petticrew et al., 2015; Skivington et al., 2021). However, none of the most influential definitions of complex interventions clearly defines what a component is. The only definition of component we are aware of is offered by iCAT_SR (Lewin et al., 2017), where a component is defined as a “discrete, active element of the intervention that could be implemented independently of other elements” (op. cit., p. 3). Based on this definition, each behavior or action (BeA) promoted by the intervention in its recipients could be considered the atomic constituent of an intervention. Each BeA may have specific implementation characteristics, which may at the same time have a verifiable effect on one or more outcomes, so that its contribution, whether direct, moderated or mediated, to the change pursued by the intervention can be better identified in a detailed and replicable report.

While the presence of multiple components and the relations that may derive from them are the first source of complexity, the second source arises from the system (e.g., an education or healthcare system, or a company) in which the intervention operates (Hawe, Shiell & Riley, 2009; Rutter et al., 2017). A system may be a multidimensional entity composed of multiple organization layers, each hosting independent yet interacting processes. When complexity is examined at the system level, an intervention can be considered an event expected to contribute to change at one or more levels and/or categories of recipients (e.g., users, professionals, managers, policymakers). Therefore, the number of levels and categories of recipients targeted by the intervention is another important factor contributing to its complexity.

Even though it is unreasonable to implement an intervention for which there is no empirical evidence of its efficacy in achieving the desired outcomes, it is also not justifiable to implement it if the report of their development does not contain the necessary information to do so in the precise manner intended by its authors. In order to improve the reporting of interventions, the MRC (Skivington et al., 2021) encourages the publication of their development separately from the assessment of its results (such as efficacy, effectiveness, efficiency or feasibility). It is to be seen the impact of this recommendation on the literature reporting interventions.

In this study, our aim was to examine the salient features of intervention reports and to explore whether some of these features are related to their quality. To ensure certain degree of homogeneity and according to our expertise, we focused on psychological interventions. According to Munn (2018), a scoping review represents the most appropriate type of systematized review to address such exploratory objectives. Specifically, our scoping review aimed to:

a. Describe the most common features of the intervention reports.
b. Analyze the replicability of the interventions.
c. Explore the presence of elements that characterize complex interventions and whether they are related to the quality and replicability of intervention reports.
d. Analyze the relationship between the quality of the intervention report and its replicability with: 1) the objective of the intervention report (devoted exclusively to the description of its development and its implementation, or also including the evaluation of its feasibility, efficacy, effectiveness or efficiency); 2) the research field to which the intervention belongs (clinical, public health, social and organizational, or education); and 3) the use of reporting tools or guidelines.

## Method

This review was conducted following the Preferred Reporting Items for Systematic Reviews and Meta-Analyses extension for Scoping Reviews (PRISMA-ScR) (Tricco et al., 2018). The protocol was prospectively registered at INPLASY (Vives et al., 2025b).

In accordance with the principles of open science, all datasets and supplementary information referenced in the following sections are publicly available in the CORA_RDR public repository and can be accessed at https://doi.org/10.34810/data2792. The PRISMA-ScR checklist applied can be consulted in <u>S1_PRISMA-ScR</u>.

### Search strategy

The search was conducted in PsycInfo (Proquest) and Web of Science (Clarivate), with the aim of collecting data from a representative sample of recently published studies focusing on reporting on psychological interventions in the four application domains: clinical, public health, social and organizational, and educational. The general search structure ((“intervention*” OR “treatment*” OR “therap*” OR “program*”) NEAR/5 (“develop*” OR “protocol” OR “feasibility” OR “effica*” OR “effectiv*”)) was adapted to accommodate specific descriptors of each of the four psychological research fields (see full search syntax in Appendix A).

### Eligibility criteria

Eligible studies were those:

1. Describing the development or implementation of psychological interventions, whether they also included empirical assessments of their feasibility, efficacy, effectiveness, or efficiency.
2. Providing information about the implementation process, even if parts of this process referenced previous publications (e.g., a previous protocol, a previously published manual, etc.).
3. Whose reported interventions belonged to the four research fields of clinical, public health, social and organizational, or educational.
4. Published between 2014 and 2024, in order to focus this study on the recent research.

### Selection process

A stratified random selection of 200 studies from each of the four target research fields was made, followed by a random sequential selection of 21 studies, one of which was used to perform the pilot test of data extraction, and the remaining 20 to extract the information of interest for our objectives, including a total of 80 studies in the review. Two reviewers (JML, JV) independently assessed the inclusion criteria based on the titles and abstracts of the studies and made the final selection from the full text. Discrepancies during this selection process were resolved by consensus between the two reviewers, with the participation of a third reviewer specialist in the field of intervention (AC, SL, JM/MFR, CM/EL/PV) when necessary. The degree of agreement between the raters was calculated using the Kappa index.

### Data collection

References identified by the search strategy were entered into Zotero bibliographic software, and duplicates were removed automatically.

Two authors (JML, JV) generated an initial list of items based on the review of: a) publications focused on establishing the characteristics that determine the complexity of interventions (Craig et al., 2008; Guise et al., 2017a; Rutter et al., 2017; Tanner-Smith & Grant, 2018; Skivington et al., 2021); and b) tools and guides to assess the quality of intervention reports, with particular attention to those that consider the complexity of interventions and incorporate guidelines or elements in their proposals aimed at capturing the characteristics of the complexity of the interventions. The tools that served as the basis for writing the items were SPIRIT (*Standard Protocol Items: Recommendations for Interventional Trials*) (Chan et al., 2013; Hopewell et al., 2022), iCAT-SR (*Complexity Assessment Tool for Systematic Reviews*) (Lewin et al., 2017), Consort-SPI (Montgomery et al., 2018), CReDECI 2 (*Criteria for Reporting the Development and Evaluation of Complex Interventions in healthcare*) (Möhler, Köpke & Meyer, 2015; Tanner-Smith & Grant, 2018), RIPI-f (*Reporting Integrity of Psychological Interventions delivered face-to-face*) (Lopez-Alcalde et al., 2022), StaRI (*Standards for Reporting Implementation Studies*) (Pinnock et al., 2015, 2017), TIDieR (*Template for Intervention Description and Replication checklist*) (Hoffmann et al., 2014) and TREND (*Transparent Reporting of Evaluations with Non-Randomized designs*) (Des Jarlais, Lyles & Crepaz, 2004).

The same authors developed a second version of the items to be used for data extraction, this time including response options. A two-round Delphi survey with all the research team (AC, CM, EL, JM, JML, JV, MR, PV, SL) was conducted using this second version as the starting point. The results of the Delphi allowed the development of a third version of the data extraction spreadsheet. Then, all research team members participated in the pilot test, which involved applying this third version to one published study from each of the four research areas. The results of a pilot test, together with the research team’s comments, led to modifications and/or additions to items and response options that were deemed necessary, resulting in the final version of the data extraction spreadsheet (see <u>S3_Data_Extraction_Spreadsheet</u>) with response options protected by drop-down lists and checkboxes (see Fig. 1a and Fig. 1b).

**Figure 1a.**
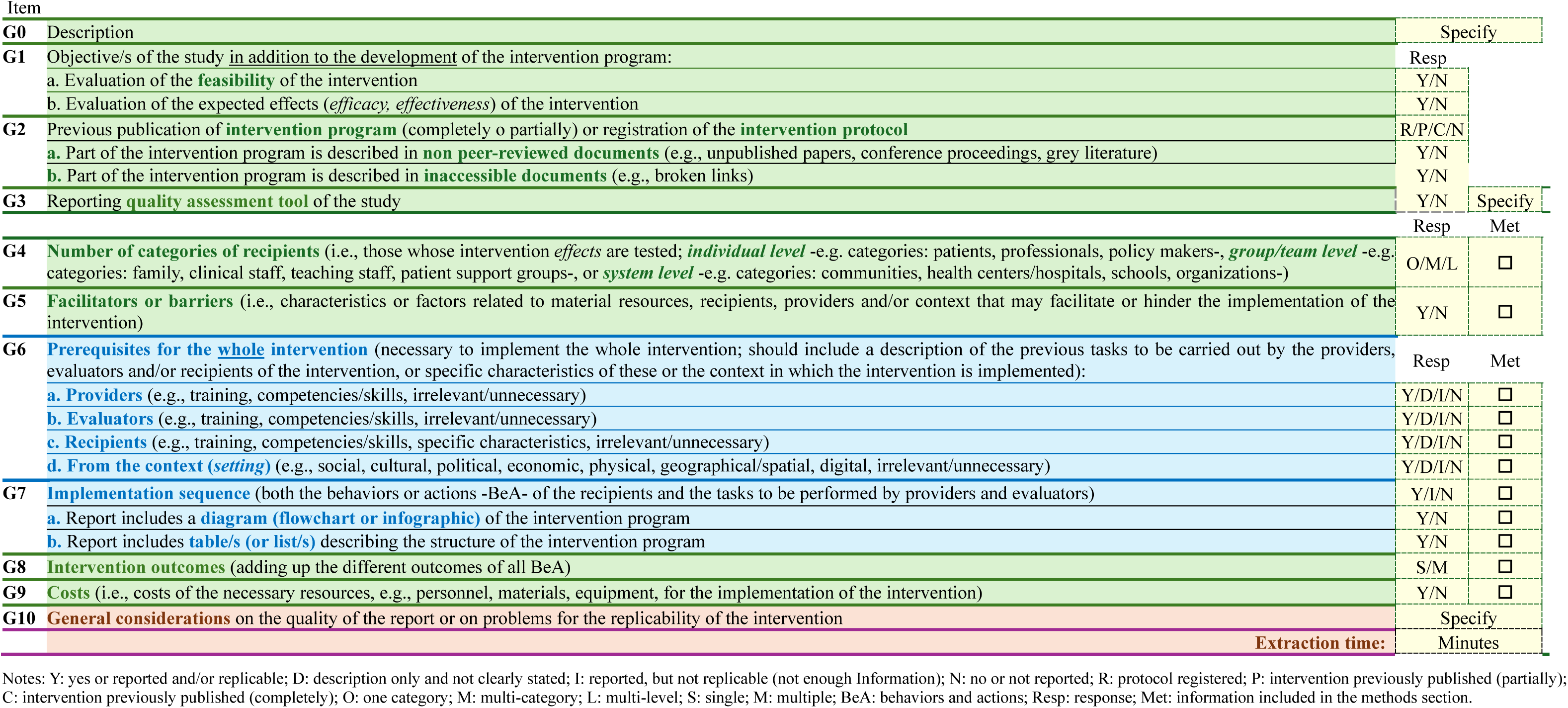
Data extraction. Report on the general characteristics of the intervention.

**Figure 1b.**
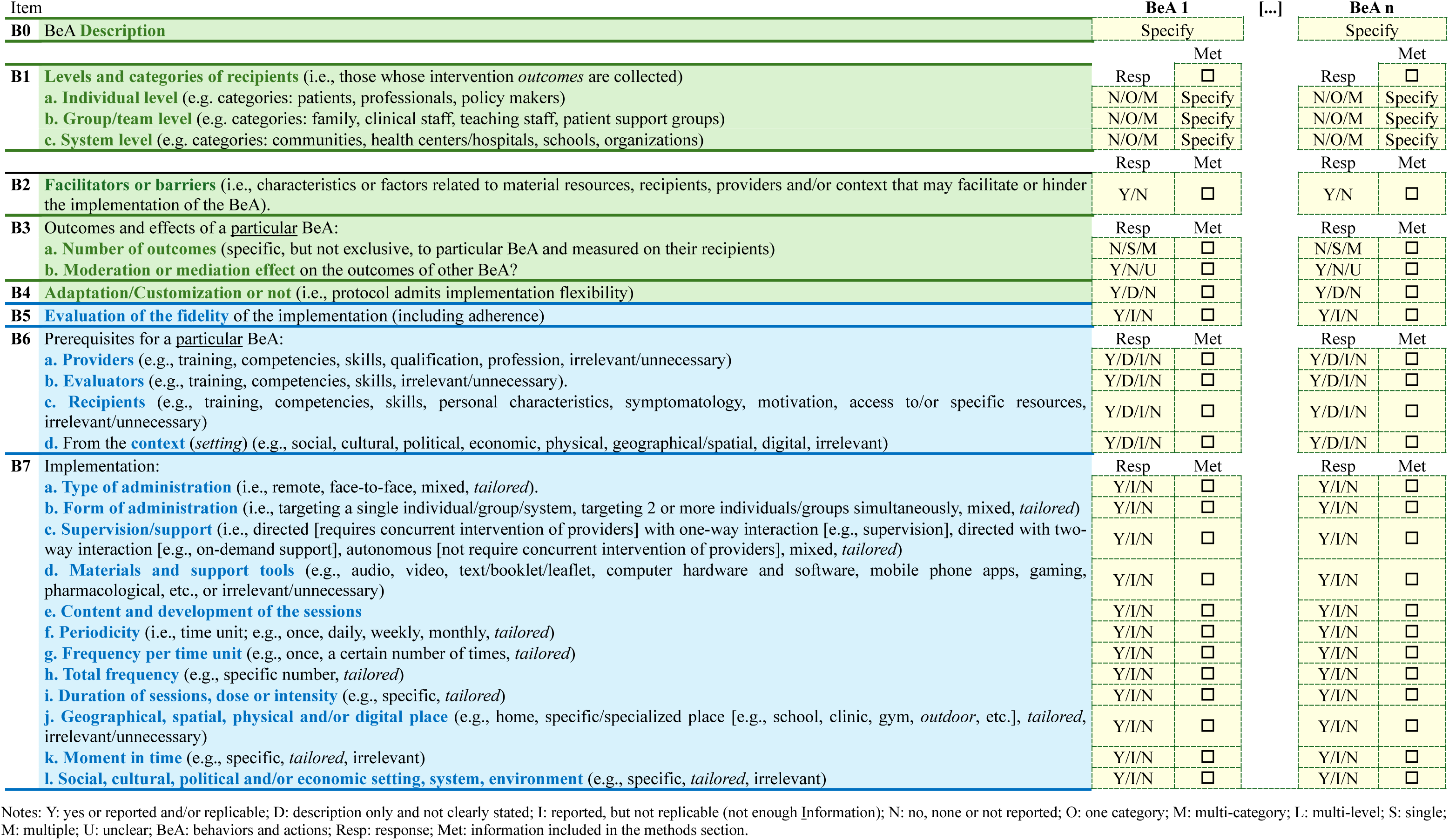
Data extraction. Report of each of the recipients’ behaviors or actions (BeA).

Although most elements of the data extraction spreadsheet are relatively straightforward, certain items warrant additional clarification. Items were categorized as green, which assessed characteristics of the intervention not directly related to its implementation, or blue, which evaluated whether the information reported on implementation was sufficient to enable accurate and unambiguous replication. While both green and blue items shared response options such as Yes and No, their interpretation differed: a Yes response for a green item indicated that the statement was factually true (e.g., a Yes for Item G1a signified that at least one study objective involved assessing the feasibility of the intervention), whereas a Yes response for a blue item signified that the reported information was replicable (e.g., a Yes for Item G7 indicated that the implementation sequence could be reproduced).

All items referring to information that should be reported in the Methods section included a checkbox (“Met”), which was ticked if that information was indeed reported in that section.

Items were organized into two spreadsheets: one containing items that pertained to the whole intervention, and another containing the items that affected each of the BeA of the intervention’s target recipients. Items G6 and B6 include a “D” response option (“description only and not clearly stated”), which is absent in the other items. These items aim to capture information about prerequisites related to providers, evaluators, recipients, and contextual factors. During the pilot test, it was observed that some studies described the characteristics of the sample (participants), the professionals who administered or measured the results, or the characteristics of the intervention’s implementation context, but did not specify whether this information referred to prerequisites for implementing the intervention or was merely a description of how the study was conducted. For these cases, response option “D” was considered.

The data extraction process was conducted similarly to the study selection process. Two reviewers performed independently the data extraction. Discrepancies during this process were resolved by consensus between the two reviewers, with the participation of a third reviewer specialist in the intervention field (AC, SL, JM/MFR, CM/EL/PV) when necessary. Prior to data extraction, the two raters identified and agreed on the BeA of each study according to the definition provided in the introduction: any constituent of an intervention with specific implementation characteristics that might also have a verifiable effect on one or more intervention outcomes was considered a potential BeA.

### Data analysis

A narrative synthesis of the results was carried out using descriptive statistical tables and bar charts. The analyses were organized according to the specific objectives set for the scoping review. Thus, a first descriptive analysis of the general characteristics of the interventions and their BeA was carried out, and the possible differences between the four research fields were explored. Secondly, possible differences in the quality of reporting (replicability) of items related to the implementation of the intervention were evaluated due to: a) the research field of the intervention; b) the use by the study authors of guidelines for writing the report; and c) the characteristics of the intervention related to its complexity, such as the number of BeA, the number of categories and levels of the intervention recipients, and the level of adaptability or customization of the implementation reported by its authors (JAMOVI v.2.6.4).

The raw and curated data may be consulted in <u>S4_Raw_Data</u> and <u>S5_Curated_Data</u>, respectively. Data extracted from the main variables is shown in Table 1.

**Table 1.** Data extraction. Main variables.

| ID | Field | G_1 | G_2 | G_3 | G_5 | G_7 | G_9 | nBeA | G_4 | BeA_1a | BeA_1b | BeA_1c | G_6 | BeA_6 | G_8 | BeA_3a | BeA_3b | BeA_4 | BeA_5 | BeA_7 |
| --- | --- | --- | --- | --- | --- | --- | --- | --- | --- | --- | --- | --- | --- | --- | --- | --- | --- | --- | --- | --- |
| Adnan, 2023 | Soc | Fea; Eff | NP | No | No | Y | No | 2 | O | O | No | No | I | I | >1 | 0 | No | No | N | I |
| Aharonovich, 2018 | Clin | Fea; Eff | NP | No | No | Y | No | 1 | O | O | No | No | I | I | >1 | 0 | No | No | N | I |
| Ahlqvist-Björkroth, 2017 | Hea | Dev | NP | No | No | Y | No | 1 | L | M | O | No | Y | Y | >1 | 0 | No | No | N | I |
| Akkermans, 2015 | Soc | Eff | CP | No | No | Y | No | 1 | O | O | No | No | I | I | >1 | 0 | No | NCS | N | I |
| Andruschko, 2018 | Edu | Fea; Eff | NP | No | Yes | N | No | 4 | O | O | No | No | I | I | >1 | 0 | No | Yes | N | I |
| Appiah, 2022 | Hea | Fea | NP | No | Yes | Y | No | 1 | O | O | No | No | I | I | >1 | 0 | No | No | N | I |
| Berman, 2016 | Clin | Dev | NP | No | No | Y | No | 1 | O | O | No | No | N | N | >1 | >1 | No | NCS | N | Y |
| Bork, 2020 | Edu | Eff | NP | No | No | Y | No | 2 | O | O | No | No | N | N | >1 | 0 | No | No | N | I |
| Boudreaux, 2015 | Hea | Fea; Eff | NP | No | No | Y | No | 1 | O | O | No | No | I | I | >1 | >1 | No | NCS | Y | I |
| Caporale-Berkowitz, 2021 | Soc | Eff | CP | No | No | Y | No | 2 | O | O | No | No | N | N | >1 | 0 | No | NCS | N | I |
| Capps, 2024 | Edu | Fea; Eff | NP | Yes | Yes | Y | No | 4 | M | M | No | No | I | Y | >1 | >1 | No | Yes | Y | Y |
| Christensen, 2014 | Hea | Eff | NP | No | No | Y | No | 1 | O | O | No | No | I | I | >1 | >1 | No | NCS | N | I |
| Chu, 2015 | Edu | Fea; Eff | CP | No | No | Y | No | 2 | L | O | O | No | I | I | >1 | 0 | No | Yes | Y | Y |
| Cohrs, 2019 | Edu | Eff | NP | No | No | Y | No | 2 | O | O | No | No | I | I | >1 | 0 | No | NCS | I | I |
| Coker, 2019 | Soc | Dev | NP | No | No | N | No | 1 | O | O | No | No | N | N | >1 | 0 | No | No | N | I |
| Drake, 2024 | Hea | Eff | NP | No | No | Y | No | 1 | O | O | No | No | N | N | >1 | 0 | No | NCS | N | I |
| Eklund, 2021 | Edu | Fea; Eff | PP | No | No | Y | No | 2 | O | O | No | No | I | I | >1 | 0 | No | NCS | I | I |
| Elderson-Van Duin, 2023 | Edu | Eff | NP | No | No | Y | No | 3 | O | No | O | No | I | I | >1 | 0 | Yes | Yes | N | I |
| Emerson, 2017 | Edu | Fea; Eff | NP | No | Yes | I | No | 3 | O | O | No | No | I | I | >1 | 0 | No | Yes | N | I |
| Erturk Kilic, 2023 | Soc | Eff | NP | No | No | Y | No | 1 | O | O | No | No | I | I | >1 | >1 | No | NCS | N | I |
| Facchini, 2016 | Hea | Fea | NP | No | Yes | Y | Yes | 5 | M | O | O | No | Y | I | >1 | 0 | Yes | Yes | N | Y |
| Faretta, 2016 | Clin | Dev | CP | No | No | Y | No | 1 | O | O | No | No | I | I | >1 | >1 | No | No | N | I |
| Farrand, 2017 | Clin | Dev | NP | No | No | Y | No | 3 | O | O | No | No | Y | Y | >1 | >1 | No | Yes | N | Y |
| Ferolino, 2024 | Soc | Fea; Eff | PP | No | No | Y | No | 4 | M | O | O | No | I | I | >1 | 0 | Unclear | No | N | I |
| Fletcher, 2024 | Soc | Dev | NP | No | No | N | No | 2 | O | O | No | No | Y | Y | >1 | 0 | No | No | N | I |
| Fung, 2018 | Hea | Eff | NP | No | No | N | No | 4 | O | O | No | No | I | I | >1 | 0 | No | Yes | N | I |
| Georgiou, 2021 | Soc | Eff | NP | No | No | I | No | 1 | O | O | No | No | I | I | >1 | 0 | No | No | N | I |
| Goldenthal, 2024 | Edu | Fea | NP | No | Yes | Y | No | 4 | L | O | No | No | I | I | >1 | 0 | Yes | No | N | I |
| Hettiarachchi, 2022 | Edu | Eff | NP | No | Yes | Y | No | 3 | O | O | No | No | I | I | 1 | 0 | Unclear | Yes | N | I |
| Huang, 2016 | Hea | Fea | PP | No | Yes | Y | No | 3 | O | O | No | No | I | I | >1 | 0 | No | No | Y | I |
| Hussong, 2024 | Hea | Fea; Eff | NP | No | No | Y | No | 3 | O | O | No | No | I | I | >1 | 0 | Unclear | Yes | I | I |
| James, 2020 | Soc | Fea | NP | No | No | Y | No | 7 | O | O | No | No | I | I | >1 | 0 | Yes | Yes | N | I |
| Janka, 2022 | Soc | Dev | Pr | No | No | Y | No | 3 | O | O | No | No | Y | Y | >1 | 0 | No | No | Y | I |
| Jarero, 2016 | Clin | Fea | PP | No | No | Y | No | 8 | O | O | No | No | I | I | 1 | 0 | Yes | Yes | N | I |
| Jiwani, 2024 | Hea | Eff | NP | No | No | N | No | 4 | O | O | No | No | Y | Y | 1 | 0 | Unclear | Yes | N | I |
| Johnsen, 2024 | Edu | Eff | Pr | No | No | Y | No | 5 | M | O | O | No | Y | Y | >1 | 0 | Yes | Yes | I | I |
| Kahler, 2014 | Hea | Fea; Eff | PP | No | No | Y | No | 8 | O | O | No | No | I | Y | >1 | 0 | No | Yes | Y | I |
| Kelly, 2024 | Soc | Dev | Pr | Yes | No | N | No | 4 | O | O | No | No | Y | Y | >1 | >1 | No | Yes | I | I |
| Kemer, 2022 | Edu | Eff | NP | No | No | Y | No | 2 | O | O | No | No | Y | Y | >1 | >1 | No | No | N | Y |
| Kjellström, 2022 | Soc | Eff | Pr | Yes | No | I | No | 3 | M | M | O | No | I | Y | >1 | >1 | No | No | N | I |
| Knaevelsrud, 2017 | Hea | Fea; Eff | Pr | No | No | I | No | 3 | O | O | No | No | I | Y | >1 | >1 | No | No | Y | I |
| Kokubo, 2023 | Soc | Eff | NP | No | No | I | No | 4 | O | O | O | No | I | Y | >1 | >1 | No | No | N | I |
| Komischke-Konnerup, 2023 | Clin | Fea; Eff | NP | No | No | Y | No | 3 | O | O | No | No | I | I | >1 | 0 | Yes | Yes | N | I |
| Kopelman-Rubin, 2019 | Clin | Fea; Eff | NP | No | No | Y | No | 4 | O | O | No | No | Y | Y | >1 | 0 | Yes | Yes | Y | I |
| Koponen, 2021 | Edu | Fea; Eff | PP | No | No | I | No | 5 | O | O | No | No | I | Y | >1 | >1 | No | Yes | Y | I |
| Kraiss, 2018 | Clin | Dev | Pr | No | No | Y | No | 4 | O | O | No | No | I | I | >1 | 0 | Yes | No | N | I |
| Kulkarni, 2014 | Edu | Eff | NP | No | No | Y | No | 3 | O | O | No | No | Y | Y | >1 | >1 | No | NCS | N | Y |
| Kuschner, 2017 | Clin | Fea | NP | No | Yes | Y | No | 4 | L | O | O | No | I | Y | >1 | 0 | No | No | N | Y |
| Kwan, 2023 | Soc | Fea; Eff | NP | Yes | Yes | Y | No | 2 | O | O | No | No | I | Y | >1 | >1 | No | No | Y | I |
| La Greca, 2016 | Hea | Fea; Eff | PP | No | Yes | I | No | 2 | O | O | O | No | I | Y | >1 | >1 | No | NCS | Y | I |
| Lovett, 2021 | Edu | Fea; Eff | PP | No | Yes | Y | No | 3 | L | No | O | O | Y | Y | >1 | >1 | No | Yes | Y | I |
| Maj, 2024 | Clin | Eff | Pr | No | Yes | Y | No | 1 | O | O | No | No | I | I | >1 | >1 | No | Yes | Y | I |
| Matsuno, 2021 | Hea | Fea; Eff | NP | Yes | Yes | Y | No | 3 | O | O | No | No | I | Y | >1 | >1 | No | Yes | Y | I |
| Mesurado, 2019 | Hea | Eff | NP | No | No | I | No | 5 | O | O | No | No | N | Y | >1 | >1 | No | No | N | I |
| Michelon, 2024 | Hea | Fea; Eff | Pr | No | No | Y | No | 3 | O | O | No | No | I | I | >1 | >1 | No | Yes | I | I |
| Molek-Winiarska, 2024 | Soc | Eff | NP | No | Yes | I | No | 4 | O | O | No | No | I | Y | >1 | >1 | No | NCS | N | I |
| Morgan, 2014 | Clin | Fea; Eff | PP | Yes | Yes | I | No | 6 | O | O | No | No | I | Y | >1 | >1 | No | No | N | I |
| Motlova, 2021 | Edu | Eff | NP | No | No | Y | No | 1 | O | O | No | No | I | I | >1 | >1 | No | No | N | I |
| Newins, 2021 | Soc | Fea; Eff | NP | No | No | Y | No | 1 | O | O | No | No | I | I | >1 | >1 | No | No | Y | I |
| Nozawa, 2022 | Edu | Fea; Eff | Pr | Yes | No | Y | No | 1 | O | O | No | No | I | I | >1 | >1 | No | No | N | I |
| O'Garro, 2023 | Clin | Eff | PP | No | Yes | I | No | 2 | O | O | No | No | I | Y | >1 | >1 | No | Yes | N | I |
| Oldershaw, 2024 | Clin | Fea; Eff | NP | Yes | Yes | I | No | 5 | O | O | No | No | I | Y | >1 | >1 | No | Yes | Y | I |
| Olsson, 2024 | Soc | Fea; Eff | Pr | Yes | Yes | Y | Yes | 1 | O | O | No | No | I | I | >1 | >1 | No | Yes | Y | I |
| Otsuka, 2015 | Clin | Fea; Eff | NP | No | No | Y | No | 1 | O | O | No | No | I | I | >1 | >1 | No | No | N | I |
| Painter, 2019 | Clin | Eff | NP | No | No | Y | No | 2 | O | O | No | No | I | I | >1 | >1 | No | No | N | I |
| Pakarinen, 2018 | Hea | Eff | Pr | Yes | No | Y | No | 2 | O | O | M | No | I | I | >1 | >1 | No | No | N | I |
| Passoni, 2018 | Soc | Eff | PP | No | No | Y | No | 3 | O | O | No | No | I | I | >1 | >1 | No | No | N | I |
| Ramirez, 2021 | Clin | Eff | NP | No | Yes | Y | No | 3 | O | O | No | No | I | I | >1 | >1 | No | No | N | I |
| Rodrigo, 2022 | Clin | Eff | CP | No | No | Y | No | 1 | O | O | No | No | I | I | >1 | >1 | No | No | N | I |
| Saffaran, 2022 | Hea | Fea; Eff | NP | No | No | Y | No | 2 | O | O | No | No | I | I | >1 | >1 | No | No | Y | I |
| Saulnier, 2024 | Clin | Fea; Eff | Pr | No | No | Y | No | 2 | O | O | No | No | I | I | >1 | >1 | No | No | Y | I |
| Schneider, 2016 | Soc | Fea; Eff | NP | No | No | I | Yes | 1 | O | O | No | No | I | I | >1 | >1 | No | No | Y | I |
| Sherwood, 2023 | Hea | Fea; Eff | CP | No | No | Y | No | 2 | O | O | No | No | I | I | >1 | >1 | No | Yes | Y | Y |
| Tran, 2020 | Edu | Fea; Eff | Pr | No | No | I | No | 1 | O | O | No | No | I | I | >1 | >1 | No | No | N | Y |
| Walker, 2014 | Hea | Eff | NP | No | No | Y | No | 1 | O | O | No | No | I | I | >1 | >1 | No | No | N | Y |
| Ward, 2020 | Soc | Eff | NP | No | No | Y | No | 2 | O | O | No | No | I | I | >1 | >1 | No | No | N | I |
| Wasik, 2020 | Edu | Fea; Eff | NP | No | No | Y | No | 3 | O | O | No | No | I | I | >1 | >1 | No | No | Y | I |
| Weigel, 2015 | Edu | Eff | Pr | Yes | No | Y | No | 1 | O | O | No | No | I | I | >1 | >1 | No | No | N | I |
| Zajonz, 2024 | Clin | Eff | NP | No | No | N | No | 1 | O | O | No | No | I | I | >1 | 0 | No | No | N | I |
| Zhang, 2014 | Clin | Eff | NP | No | No | Y | No | 3 | O | O | No | No | I | I | >1 | 0 | No | Yes | N | I |

Note 1. Table Columns. Field: Research field; G\_1: Objectives; G\_2: Previous publication of intervention program or protocol; G\_3: Reporting quality assessment tool; G\_5: Facilitators or barriers; G\_7: Implementation sequence; G\_9: Costs; nBeA: Number of BeA; G\_4: Number of categories of recipients; BeA\_1a: Levels & categories of recipients: Individual level; BeA\_1b: Levels & categories of recipients: Group/team level; BeA\_1c: Levels & categories of recipients: System level; G\_6: Prerequisites of the whole intervention: Global; BeA\_6: Prerequisites of the particular BeA: Global; G\_8: Intervention outcomes; BeA\_3a: Number of outcomes/effects; BeA\_3b: Moderation or mediation effect on the outcomes of other BeA; BeA\_4: Adaptation/customization; BeA\_5: Evaluation of the fidelity (including adherence); BeA\_7 Implementation: Global
Note 2: Abbreviations: Clin: Clinical; Edu: Education; Hea:Health; Soc: Social; Eff: Effectiveness; Fea: Feasibility; Dev: Development only; CP: Intervention previously published (completely); NP: Without prior publication or registration; PP: Intervention previously published (partially); Pr: Protocol registered; L: Multi-level; M: Multi-category; O: One-category; D: Description only, no prerequisite stated; I: Reported, but not replicable (missing info); N: Not reported; Y: Reported and/or replicable; NCS: Not clearly stated

### Ethics statement

This scoping review does not require approval from an ethics committee, as no data was collected directly from humans or animals.

## Results

### Search results

Figure 2 shows the results of the search strategy, reported according to the PRISMA flowchart (Page et al., 2021). Finally, 84 studies were selected, 4 were used for testing the extraction procedure, and the remaining 80 were included in the review. The interrater agreement for abstract and title selection was 100%, so no kappa coefficient needed to be calculated, whereas the interrater agreement for full text selection was kappa k=0.78 (CI95%: 0.59-0.96). After discussion to solve the discrepancies, the interrater agreement percentage for full text selection was 100%.

**Figure 2.**
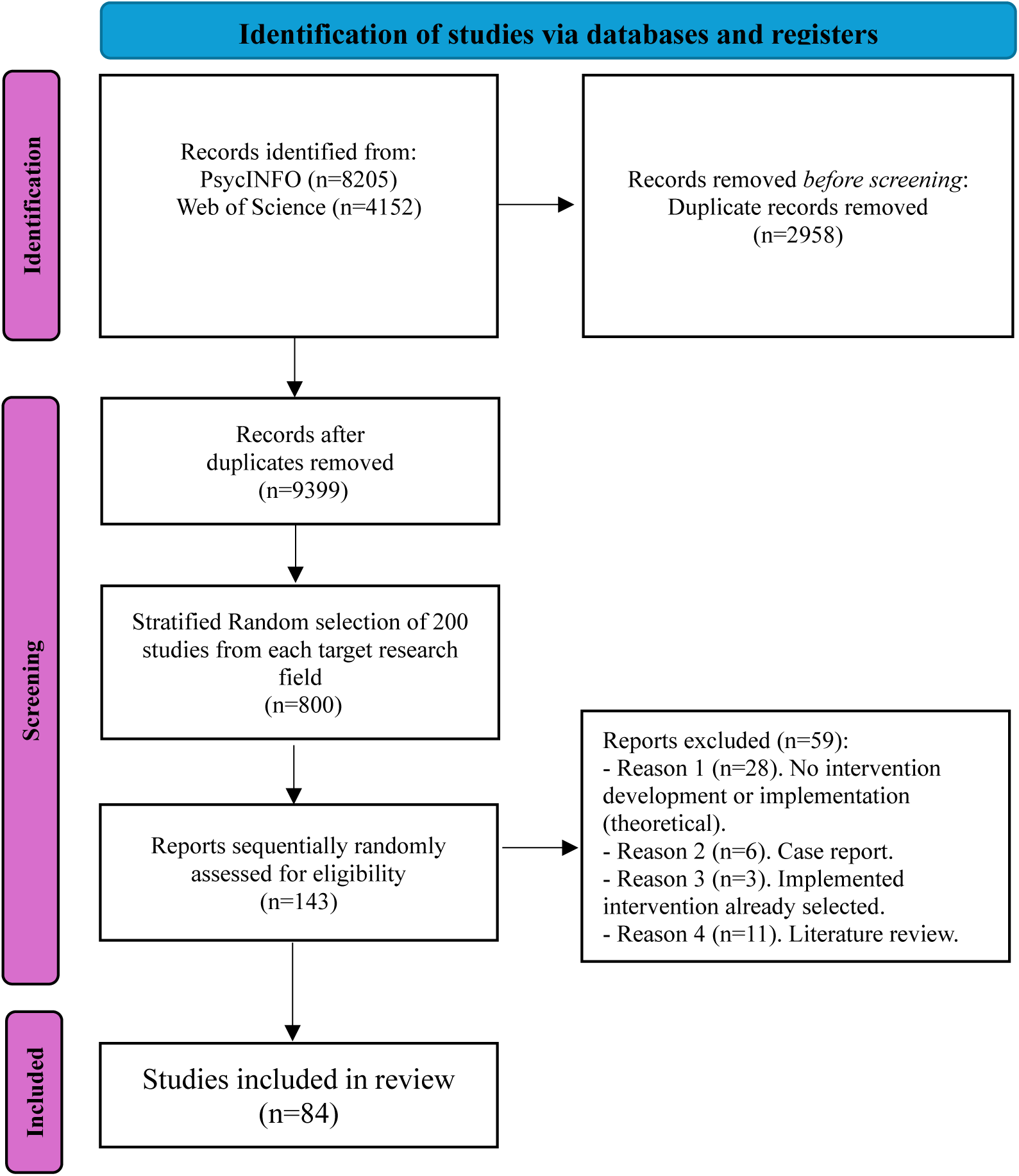
PRISMA flow diagram of the selection process.

### Characteristics of the whole interventions

Table 2 and Figure 3 summarize the key characteristics of intervention programs in four research fields (clinical, health, social, and education). The analysis included chi-square tests to assess differences between fields. Below, we first describe the overall patterns observed for each characteristic across all fields combined, followed by a comparison across the individual fields.

**Figure 3.**
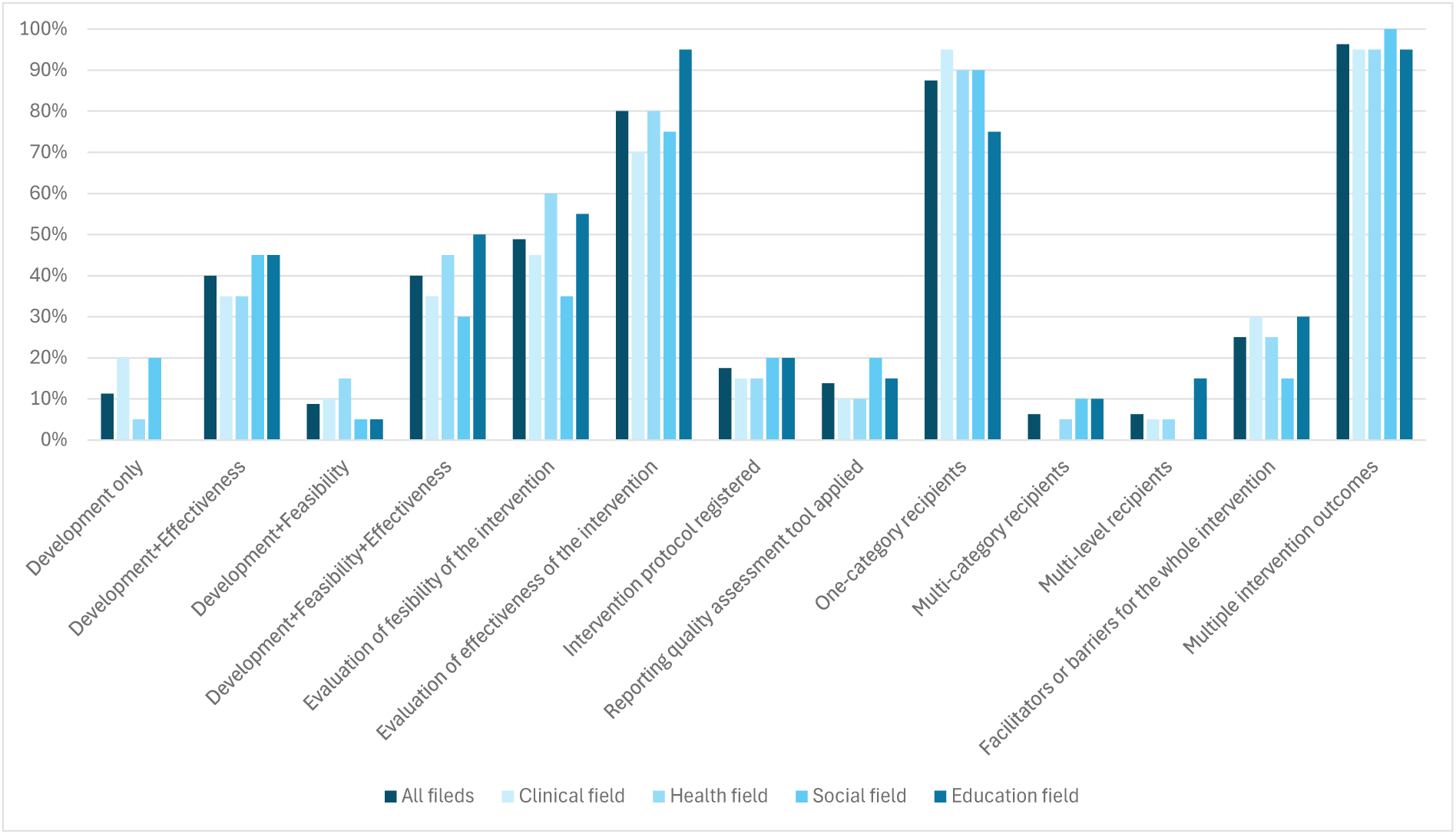
Characteristics of the whole interventions.

**Table 2.**
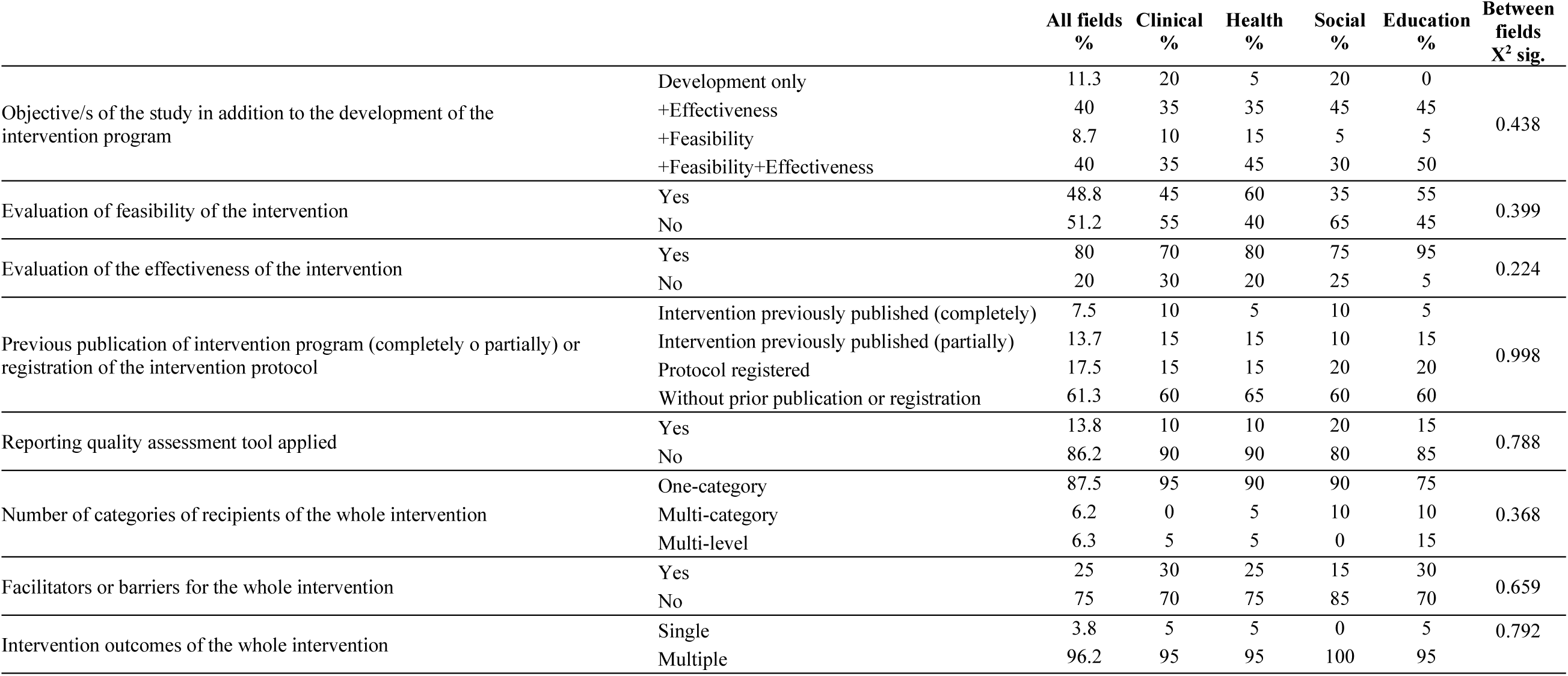
Characteristics of the whole intervention by field.

Across all fields, only 11.3% of studies focused exclusively on the development of the intervention program. Most publications combined development with an additional objective, most often the assessment of effectiveness (40%) or both feasibility and effectiveness (40%). Feasibility evaluation was reported in just under half of the studies (48.8%), while the majority (80%) assessed the effectiveness. Most studies (61.3%) did not previously publish the intervention or register their protocol, and only 13.8% applied a reporting quality assessment tool.

In terms of recipients, most interventions targeted a single category (87.5%), while a small number addressed multiple levels (6.3%). Facilitators or barriers for the intervention were reported in only 25% of cases. Finally, nearly all interventions (96.3%) included multiple outcomes.

No statistically significant differences were found across research fields, though descriptive trends emerged. Development-only objectives appeared in clinical and social research (20% each) but were absent in education, while health studies rarely stopped at development (5%). Combined evaluations of feasibility and effectiveness were most frequent in education (50%) and least in social research (30%). Feasibility assessments dominated health (60%) and education (55%) but were less common in social studies (35%). Effectiveness was almost universally assessed in education (95%) and less so in clinical research (70%).

Reporting quality tools were rarely used, ranging from 10% of clinical and health to 20% of social studies. Most interventions targeted a single recipient category, especially in clinical (95%) and health (90%), whereas education showed greater diversity, with 15% of multi-level interventions compared to 0-5% in the other fields. Finally, reporting of facilitators or barriers was highest in education and clinical (30%) and lowest in social research (15%).

### Characteristics of the intervention’s recipients’ behaviors or actions (BeA)

Table 3 and Figure 4 outline the characteristics related to the BeA, both at global level and for each of the four specific research fields that were analyzed.

**Figure 4.**
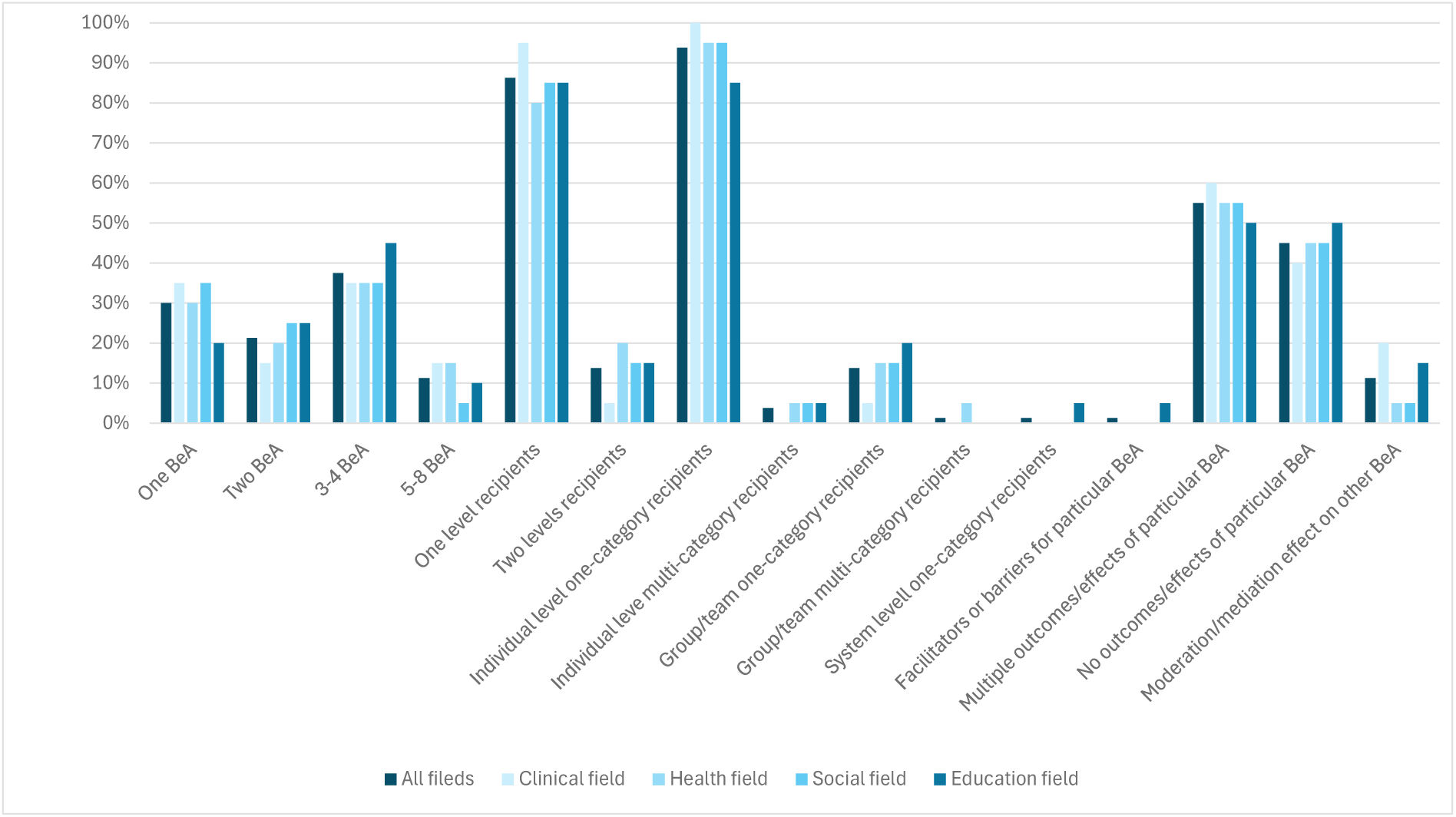
Characteristics of the intervention recipients’ behaviors or actions (BeA).

**Table 3.** Characteristics of the intervention’s recipients’ behaviors or actions (BeA) by field.

|  |  | All fields<br>% | Clinical<br>% | Health<br>% | Social<br>% | Education<br>% | Between<br>fields<br>X<sup>2</sup> sig. |
| --- | --- | --- | --- | --- | --- | --- | --- |
| Number of BeA | One BeA | 30 | 35 | 30 | 35 | 20 | 0.953 |
|  | Two BeA | 21.2 | 15 | 20 | 25 | 25 |  |
|  | 3-4 BeA | 37.5 | 35 | 35 | 35 | 45 |  |
|  | 5-8 BeA | 11.3 | 15 | 15 | 5 | 10 |  |
| Number of levels of recipients of particular BeA | One level | 86.2 | 95 | 80 | 85 | 85 | 0.572 |
|  | Two levels | 13.8 | 5 | 20 | 15 | 15 |  |
| Levels & categories of recipients of particular BeA: Individual level | One-category | 93.8 | 100 | 95 | 95 | 85 | 0.298 |
|  | Multi-category | 3.7 | 0 | 5 | 5 | 5 |  |
|  | No | 2.5 | 0 | 0 | 0 | 10 |  |
| Levels & categories of recipients of particular BeA: Group/team level | One-category | 13.7 | 5 | 15 | 15 | 20 | 0.534 |
|  | Multi-category | 1.3 | 0 | 5 | 0 | 0 |  |
|  | No | 85 | 95 | 80 | 85 | 80 |  |
| Levels & categories of recipients of particular BeA: System level | One-category | 1.3 | 0 | 0 | 0 | 5 | 0.386 |
|  | No | 98.7 | 100 | 100 | 100 | 95 |  |
| Facilitators or barriers for particular BeA | Yes | 1.2 | 0 | 0 | 0 | 5 | 0.386 |
|  | No | 98.8 | 100 | 100 | 100 | 95 |  |
| Number of outcomes/effects of particular BeA | Multiple | 55 | 60 | 55 | 55 | 50 | 0.939 |
|  | None | 45 | 40 | 45 | 45 | 50 |  |
| Moderation or mediation effect of particular BeA on the outcomes of other BeA | Yes | 11.3 | 20 | 5 | 5 | 15 | 0.523 |
|  | Unclear | 5 | 0 | 10 | 5 | 5 |  |
|  | No | 83.7 | 80 | 85 | 90 | 80 |  |

Across all fields, most interventions included more than one recipients’ BeA. While 30% addressed a single BeA and 21.3% two BeA, the largest proportion (37.5%) reported 3–4 BeA, and a smaller proportion (11.3%) addressed 5–8 BeA. Most studies described recipients’ behaviors at a single level (86.3%), 97.6% considered the individual level, 15.1% the group/team level, and only 1.3% the system level.

Facilitators or barriers for particular BeA were almost absent (1.3%). In terms of outcomes, just over half of the interventions (55%) reported multiple outcomes or effects for BeA, while 45% reported none. Interactive effects among BeA were infrequently explored: only 11.3% of studies reported moderating or mediating effects of the BeA on the outcomes of other BeA, and 5% were unclear.

No statistically significant differences were observed between fields. Nevertheless, some descriptive variations can be noted. Education showed a slightly higher proportion of interventions with 3 or 4 BeA (45%), while clinical and social fields had somewhat higher rates of single BeA (35% each). Clinical studies reported the highest proportion of single-level interventions (95%), while health interventions were slightly more likely to include two recipient levels (20%). At individual level, education showed slightly more diversity, with 10% of studies not reporting this level, while at group/team level, education (20%) and social (15%) included it slightly more often than clinical (5%). At system level, only education (5%) reported it. Finally, clinical research led in testing moderation and/or mediation effects (20%), followed by education (15%), with health and social both at 5%.

### Quality reporting of implementation characteristics of the whole interventions

Table 4 and Figure 5 present the quality reporting of implementation of the intervention studies analysed, categorized by reporting status: reported and/or replicable (RR), reported but not replicable due to missing information (RNR), and not reported (NR).

**Figure 5.**
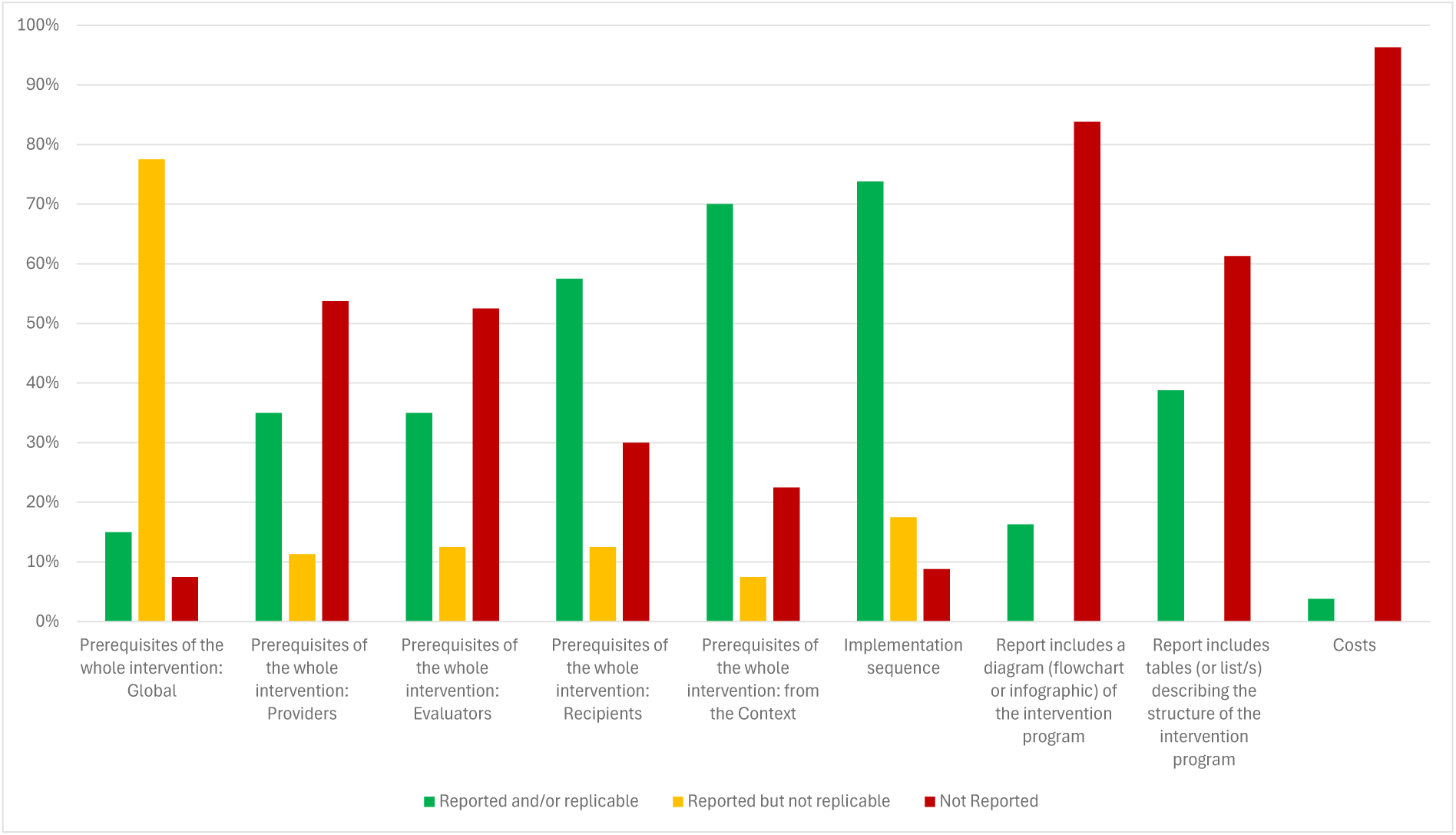
Quality reporting of implementation characteristics of the whole interventions.

**Table 4.** Quality reporting of implementation characteristics by field.

|  | All fields |  |  | Clinical field |  |  | Health field |  |  | Social field |  |  | Education field |  |  | Between fields |
| --- | --- | --- | --- | --- | --- | --- | --- | --- | --- | --- | --- | --- | --- | --- | --- | --- |
|  | RR<br>% | RNR<br>% | NR<br>% | RR<br>% | RNR<br>% | NR<br>% | RR<br>% | RNR<br>% | NR<br>% | RR<br>% | RNR<br>% | NR<br>% | RR<br>% | RNR<br>% | NR<br>% | X<sup>2</sup> sig. |
| **Implementation characteristics of the whole intervention** |  |  |  |  |  |  |  |  |  |  |  |  |  |  |  |  |
| Global prerequisites of the whole intervention | 15 | 77.5 | 7.5 | 10 | 85 | 5 | 15 | 75 | 10 | 15 | 75 | 10 | 20 | 75 | 5 | 0.958 |
| Providers' prerequisites | 35 | 11.3 | 53.7 | 40 | 5 | 55 | 50 | 0 | 50 | 20 | 30 | 50 | 30 | 10 | 60 | 0.055 |
| Evaluators' prerequisites | 35 | 12.5 | 52.5 | 20 | 15 | 65 | 45 | 0 | 55 | 35 | 25 | 40 | 40 | 10 | 50 | 0.208 |
| Recipients' prerequisites | 57.5 | 12.5 | 30 | 75 | 5 | 20 | 60 | 10 | 30 | 50 | 25 | 25 | 45 | 10 | 45 | 0.256 |
| Context prerequisites | 70 | 7.5 | 22.5 | 65 | 0 | 35 | 65 | 15 | 20 | 75 | 10 | 15 | 75 | 5 | 20 | 0.467 |
| Global prerequisites of the whole intervention or for BeA | 13.8 | 78.7 | 7.5 | 10 | 85 | 5 | 10 | 80 | 10 | 15 | 75 | 10 | 20 | 75 | 5 | 0.934 |
| Implementation sequence | 73.8 | 17.5 | 8.7 | 80 | 15 | 5 | 75 | 15 | 10 | 60 | 25 | 15 | 80 | 15 | 5 | 0.789 |
| Report includes a diagram of the intervention program | 16.2 | 0 | 83.8 | 20 | 0 | 80 | 25 | 0 | 75 | 10 | 0 | 90 | 10 | 0 | 90 | 0.479 |
| Report includes tables/lists of the structure of the intervention program | 38.8 | 0 | 61.2 | 65 | 0 | 35 | 20 | 0 | 80 | 30 | 0 | 70 | 40 | 0 | 60 | 0.024* |
| Costs | 3.7 | 0 | 96.3 | 0 | 0 | 100 | 5 | 0 | 95 | 10 | 0 | 90 | 0 | 0 | 100 | 0.283 |
| **Implementation characteristics of recipients' behaviors or actions (BeA)** |  |  |  |  |  |  |  |  |  |  |  |  |  |  |  |  |
| Adaptation/Customization or not | 35 | 15 | 50 | 40 | 5 | 55 | 40 | 20 | 40 | 15 | 20 | 65 | 45 | 15 | 40 | 0.326 |
| Evaluation of the fidelity/adherence of the implementation | 27.5 | 7.5 | 65 | 20 | 0 | 80 | 40 | 10 | 50 | 25 | 5 | 70 | 25 | 15 | 60 | 0.533 |
| Global prerequisites for particular BeA | 32.5 | 61.3 | 6.2 | 30 | 65 | 5 | 35 | 60 | 5 | 35 | 55 | 10 | 30 | 65 | 5 | 0.978 |
| Providers' prerequisites | 41.3 | 3.7 | 55 | 45 | 5 | 50 | 55 | 5 | 40 | 35 | 0 | 65 | 30 | 5 | 65 | 0.619 |
| Evaluators' prerequisites | 43.8 | 0 | 56.2 | 30 | 0 | 70 | 55 | 0 | 45 | 45 | 0 | 55 | 45 | 0 | 55 | 0.459 |
| Recipients' prerequisites | 61.2 | 2.5 | 36.3 | 75 | 5 | 20 | 60 | 5 | 35 | 60 | 0 | 40 | 50 | 0 | 50 | 0.466 |
| Context prerequisites | 82.5 | 5 | 12.5 | 85 | 0 | 15 | 85 | 5 | 10 | 75 | 15 | 10 | 85 | 0 | 15 | 0.361 |
| Global characteristics of the implementation | 13.8 | 86.2 | 0 | 15 | 85 | 0 | 15 | 85 | 0 | 0 | 100 | 0 | 25 | 75 | 0 | 0.146 |
| Type of administration (e.g., remote, face-to-face) | 73.7 | 21.3 | 5 | 80 | 15 | 5 | 70 | 30 | 0 | 60 | 25 | 15 | 85 | 15 | 0 | 0.198 |
| Form of administration (e.g., individual, group) | 80 | 16.2 | 3.8 | 90 | 5 | 5 | 80 | 15 | 5 | 65 | 30 | 5 | 85 | 15 | 0 | 0.446 |
| Supervision/support (e.g., directed, autonomous, mixed) | 72.5 | 22.5 | 5 | 80 | 15 | 5 | 75 | 25 | 0 | 65 | 25 | 10 | 70 | 25 | 5 | 0.807 |
| Materials and support tools (e.g., multimedia, text, app) | 47.5 | 36.3 | 16.2 | 60 | 20 | 20 | 50 | 45 | 5 | 40 | 35 | 25 | 40 | 45 | 15 | 0.405 |
| Content and development of the sessions | 36.3 | 55 | 8.7 | 65 | 25 | 10 | 25 | 70 | 5 | 10 | 80 | 10 | 45 | 45 | 10 | 0.011* |
| Periodicity (time unit; e.g., once, daily, weekly, monthly) | 75 | 11.3 | 13.7 | 90 | 0 | 10 | 60 | 15 | 25 | 75 | 15 | 10 | 75 | 15 | 10 | 0.354 |
| Frequency per time unit (e.g., once, a number of times) | 70 | 13.8 | 16.2 | 75 | 10 | 15 | 60 | 20 | 20 | 70 | 15 | 15 | 75 | 10 | 15 | 0.948 |
| Total frequency (e.g., specific number, tailored) | 76.2 | 12.5 | 11.3 | 90 | 0 | 10 | 70 | 15 | 15 | 75 | 15 | 10 | 70 | 20 | 10 | 0.591 |
| Duration of sessions, dose or intensity (specific, tailored) | 56.3 | 20 | 23.7 | 55 | 10 | 35 | 55 | 25 | 20 | 50 | 20 | 30 | 65 | 25 | 10 | 0.540 |
| Geographical, spatial, physical and/or digital place | 88.7 | 10 | 1.3 | 90 | 10 | 0 | 95 | 5 | 0 | 70 | 25 | 5 | 100 | 0 | 0 | 0.083 |
| Moment in time (e.g., specific, tailored, irrelevant) | 95 | 0 | 5 | 95 | 0 | 5 | 100 | 0 | 0 | 90 | 0 | 10 | 95 | 0 | 5 | 0.551 |
| Social, cultural, political and/or economic setting | 91.2 | 5 | 3.8 | 90 | 10 | 0 | 95 | 5 | 0 | 85 | 5 | 10 | 95 | 0 | 5 | 0.444 |

Notes: RR: reported and/or replicable; RNR: reported but not replicable (missing information); NR: not reported; \*: statistically significant (p<0.05)

Across fields, the reporting of implementation characteristics at the level of the whole intervention showed a mixed picture, with notable weaknesses in several areas. Global prerequisites of the intervention were seldom reported in sufficient detail: only 15% of studies provided replicable information, while most either offered incomplete descriptions (77.5%) or omitted them altogether (7.5%). This pattern was consistent across the different groups of prerequisites. Although recipients’ (57.5%) and context-related prerequisites (70%) were more frequently addressed, information on providers (35%) and evaluators (35%) was less often available in a replicable form. When prerequisites of the whole intervention and recipients’ behaviors were considered jointly, reporting was very limited, with only 13.8% adequately covered. By contrast, the implementation sequence was more consistently described: nearly three-quarters of studies (73.8%) included enough detail to allow replication. Nevertheless, structured reporting formats that could enhance clarity and replicability were underutilized. Only 16.3% of studies presented a diagram, and fewer than half (38.8%) provided tables or lists outlining the intervention’s structure. Costs were almost entirely neglected (3.8%).

No statistically significant differences were observed between fields in the percentage of characteristics well reported (see Fig. 6), except for the reporting of tables or lists of intervention structure (*p = 0.024*), which was highest in clinical (65%) and education (40%) compared to health (20%). Recipients’ prerequisites were most often reported in a replicable manner in clinical (75%) and health (60%) compared to education (45%, and providers’ prerequisites were adequately reported more often in health (50%) than in education (30%).

**Figure 6.**
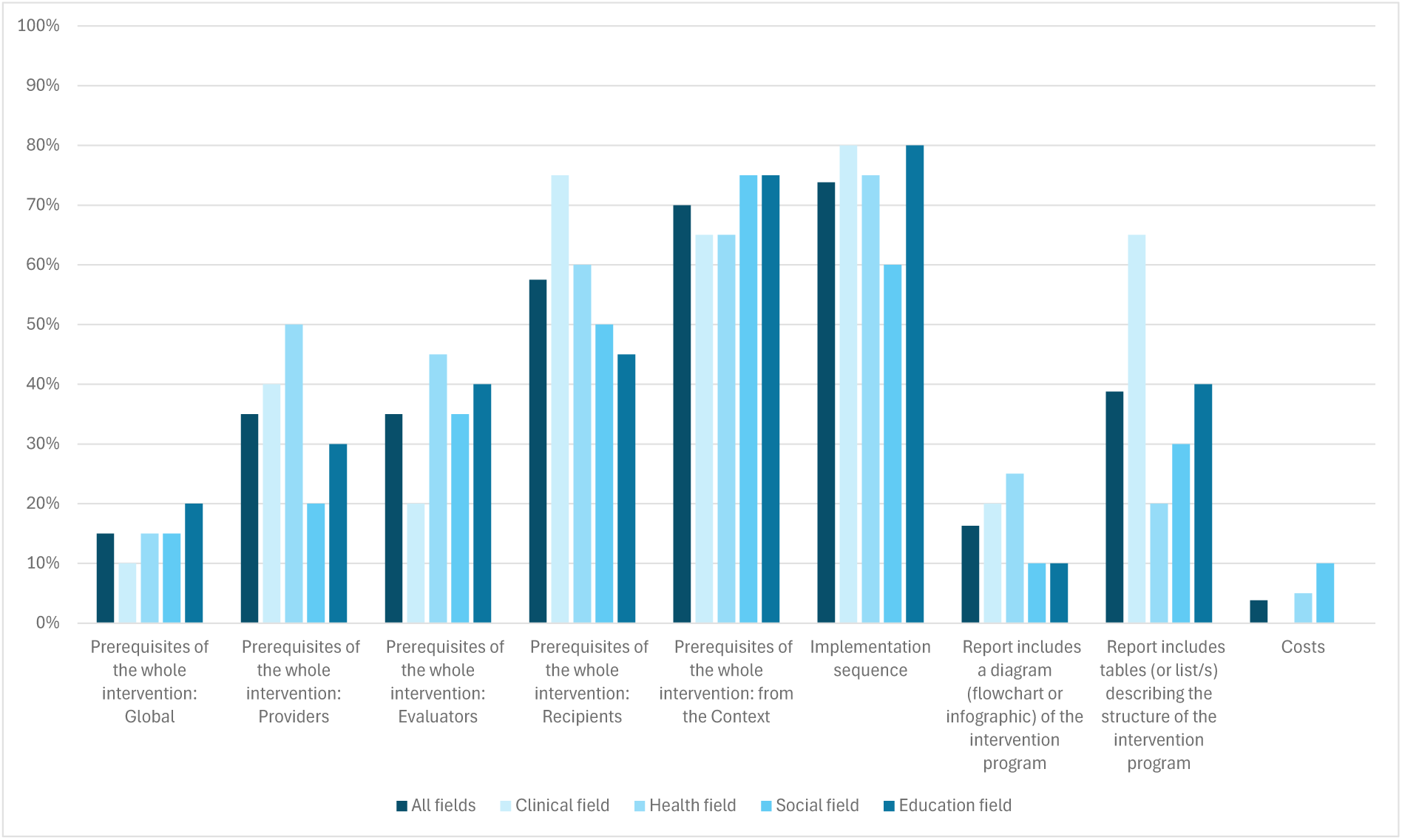
Percentage of implementation characteristics of the whole interventions reported and/or replicable by field.

Reporting of tables or lists describing the structure of the intervention program was more frequent in clinical (65%) compared to other fields (20–40%).

Finally, it is worth noting that, regarding the section of the publications in which the previously described general characteristics of the interventions were reported, these were not included in the Method section of the studies in between 14.6% (administrator prerequisites) and 37% (evaluator prerequisites) of the cases. Moreover, in 75.8% of the studies, the reporting of contextual prerequisites (setting) was also located in sections other than the Method section.

### Quality reporting of implementation characteristics of the intervention’s recipients’ behaviors or actions (BeA)

At the level of BeA, reporting quality of the implementation characteristics of the interventions also varied (Table 4 and Fig. 7). The reporting of BeA adaptation or customization—entailing whether the intervention was adaptable or customizable, and, if so, what adaptations or customizations were permitted—was done in a replicable manner in just over a third of studies (35%), with half failing to address this aspect at all. Similarly, fidelity and adherence were reported by 27.5% of studies with adequate detail and nearly two-thirds (65%) omitting this implementation feature.

**Figure 7.**
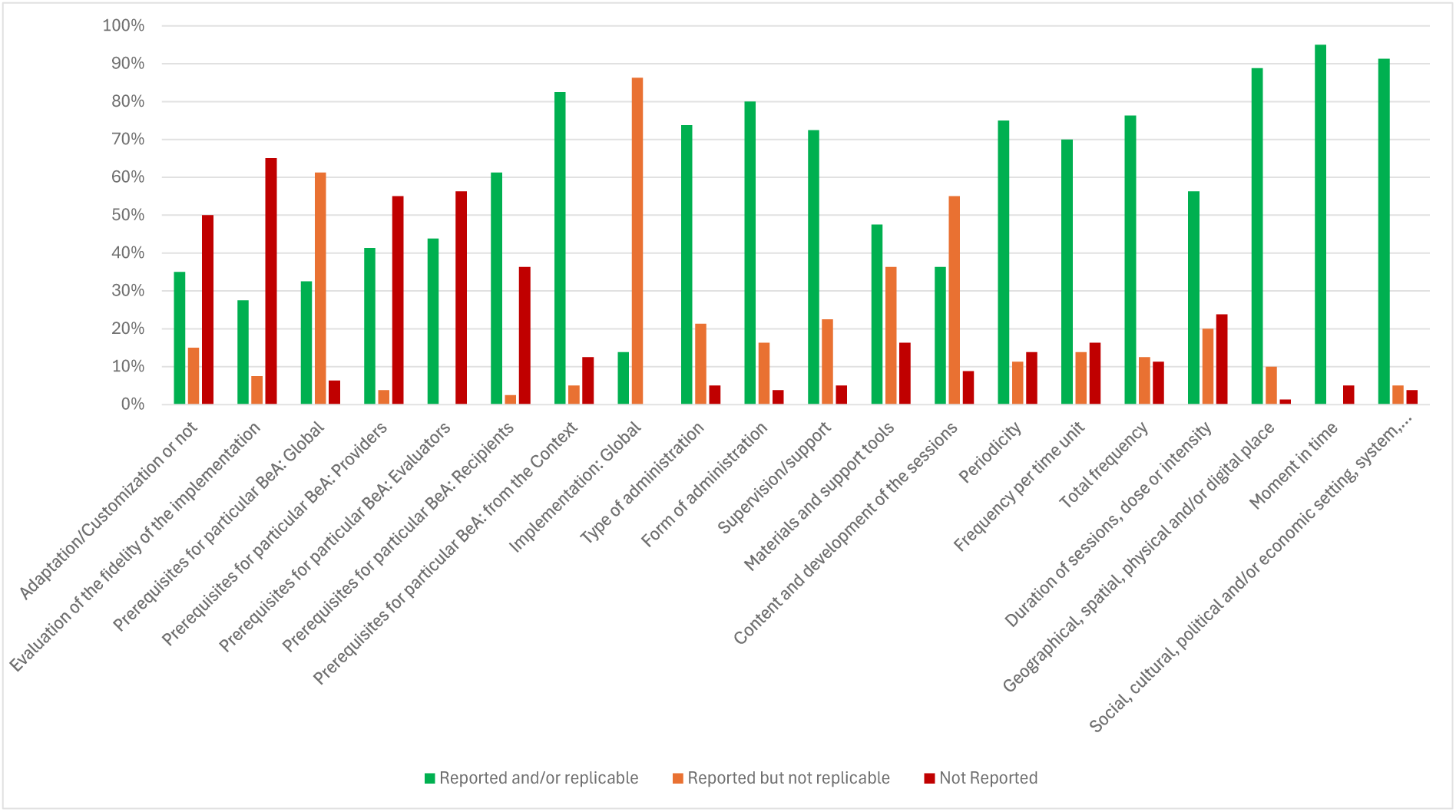
Quality reporting of implementation characteristics of the intervention’s recipients’ behaviors or actions (BeA).

Global prerequisites for recipients’ behaviours were also weakly documented: while 32.5% reported them with replicable detail, most studies (61.3%) provided incomplete information. When broken down by type of prerequisite, reporting was uneven. Recipients’ (61.3%) and context-related prerequisites (82.5%) were more frequently specified. By contrast, providers (41.3%) and evaluators (43.8%) were less consistently addressed.

In contrast, certain global characteristics of implementation were relatively comprehensively addressed. Most studies showed adequate reporting on key structural aspects, such as type (73.8%) and form of administration (80%), supervision or support (72.5%), and periodicity (75%). Frequency (70–76.3%), location (88.8%), and timing (95%) were also commonly well reported. Moreover, the social, cultural, political, and economic setting was frequently reported and/or replicable (91.3%). However, two key elements remained comparatively underreported: the content and development of sessions (36.3%) and the materials or support tools used (47.5%). The reporting of the dose or intensity of the sessions was replicable in just over half (56.3%) of the studies. Overall, the low percentage of “not reported” responses observed for most of the features analysed should be interpreted with caution, as while they indicated that these implementation features were universally addressed, this was rarely done in a fully replicable manner in light of the observed percentage of “reported but not replicable (missing information)” results.

No statistically significant differences were found across fields (Fig. 8) except for content and development of sessions (*p = 0.011*), where the replicability of the reports was greater in the clinical field (65%) and lowest in health (25%) and social (10%) fields. Adaptation/customization was reported more frequently and accurately in clinical and health fields (40% each) than in social (15%). Reporting on implementation fidelity was most adequate in health (40%) and lowest in clinical (20%) field.

**Figure 8.**
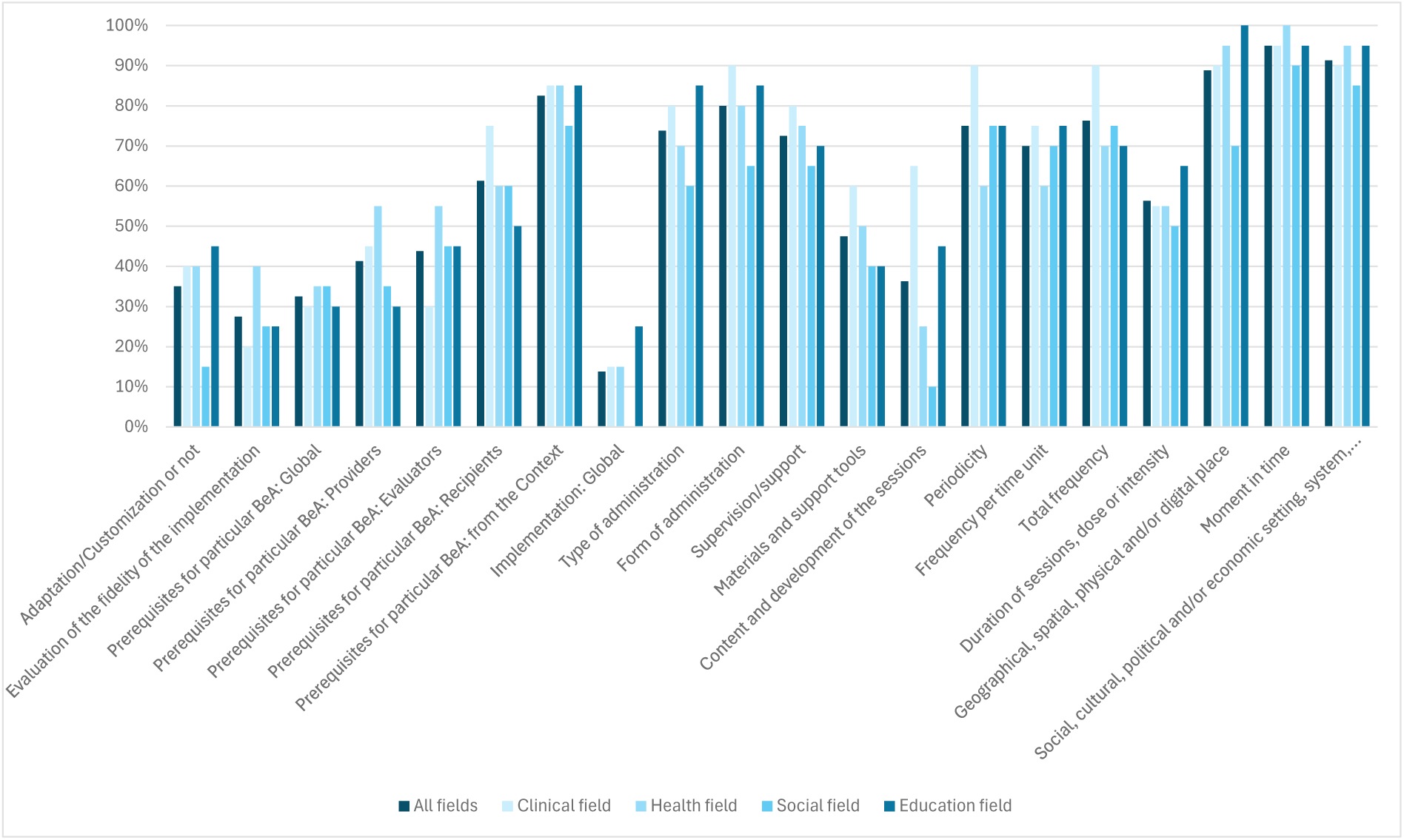
Percentage of implementation characteristics of intervention’s recipients’ behaviors or actions (BeA) reported and/or replicable by field.

Finally, it should be noted that, concerning the section of the publications in which the characteristics previously described of the BeA were reported, the four groups of implementation prerequisites were not included in the Method section in between 70% (administrator prerequisites) and 86.4% (evaluator prerequisites) of the studies. The remaining implementation characteristics of the BeA were reported outside the Method section in less than 15% of the studies, except for the materials and support tools required for BeA implementation, their potential for adaptation/customization, and the procedures used to assess implementation fidelity, which were reported outside the Method section in 37.3%, 25%, and 17.9% of the studies, respectively.

### Quality reporting of implementation characteristics of the interventions by number of intervention’s recipients’ behaviors or actions (BeA)

Table 5 and Figure 9 examine the quality of reporting for implementation characteristics in intervention studies, stratified by the number of BeA involved (1 BeA, 2 BeA, 3 BeA, 4 BeA, and 5-8 BeA). The reporting of implementation characteristics for the whole intervention varied substantially depending on the number of BeA considered. When only one BeA was addressed, replicable reporting of global prerequisites was almost absent (4.2%), and only a minority of studies included providers’ (16.7%) or evaluators’ (12.5%) prerequisites, and recipients’ prerequisites were reported in a replicable manner somewhat more frequently (25%), and context prerequisites were well documented at a relatively high level (79.2%).

**Figure 9.**
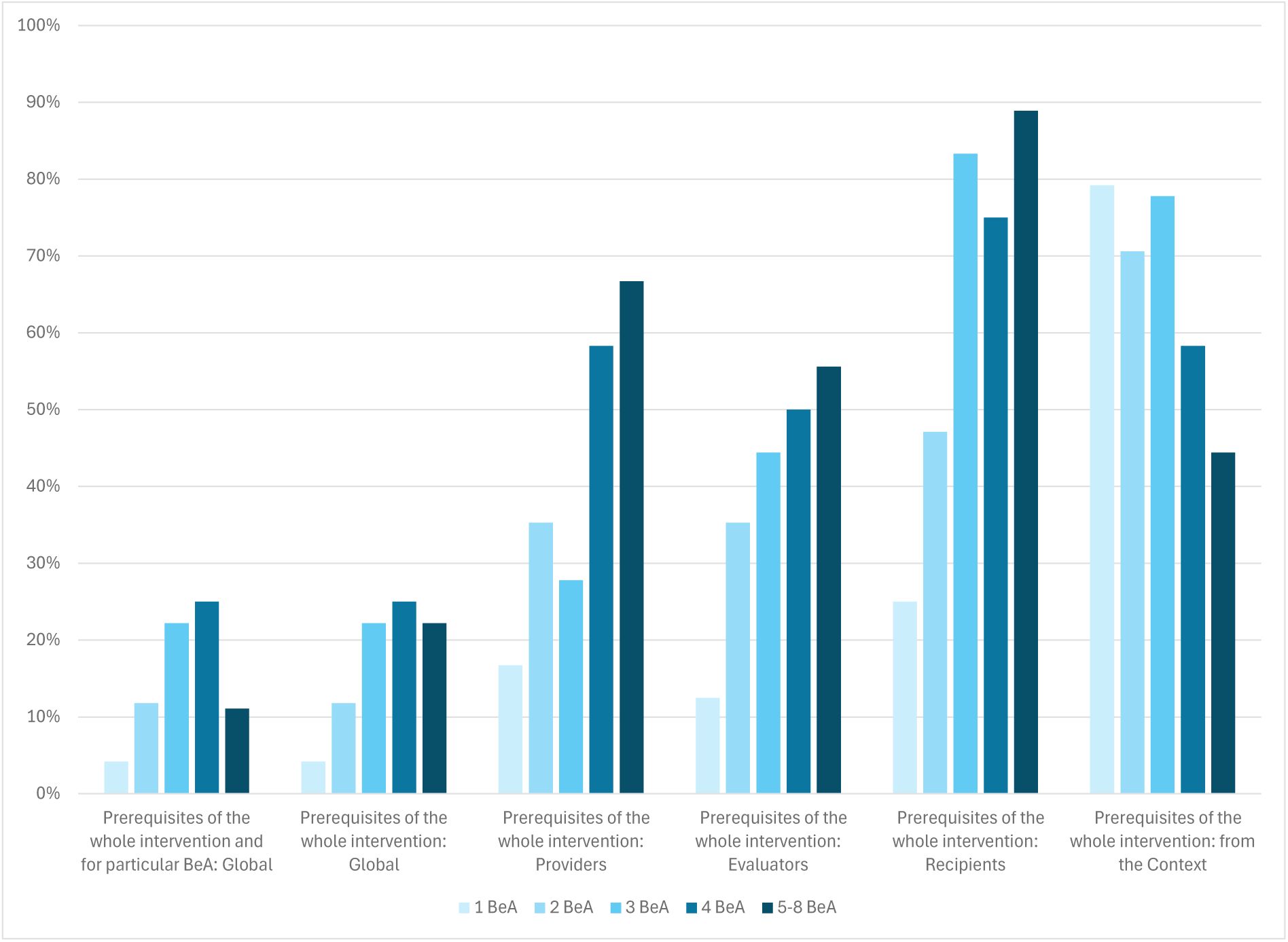
Percentage of implementation characteristics of the whole interventions reported and/or replicable by number of recipients’ behaviors or actions (BeA).

**Table 5.** Quality reporting of implementation characteristics by number of BeA.

|  | 1 BeA |  |  | 2 BeA |  |  | 3 BeA |  |  | 4 BeA |  |  | 5-8 BeA |  |  | Between fields |
| --- | --- | --- | --- | --- | --- | --- | --- | --- | --- | --- | --- | --- | --- | --- | --- | --- |
|  | RR<br>% | RNR<br>% | NR<br>% | RR<br>% | RNR<br>% | NR<br>% | RR<br>% | RNR<br>% | NR<br>% | RR<br>% | RNR<br>% | NR<br>% | RR<br>% | RNR<br>% | NR<br>% | X<sup>2</sup> sig. |
| **Implementation characteristics of the whole intervention** |  |  |  |  |  |  |  |  |  |  |  |  |  |  |  |  |
| Global prerequisites of the whole intervention | 4.2 | 83.3 | 12.5 | 11.8 | 76.4 | 11.8 | 22.2 | 77.8 | 0 | 25 | 75 | 0 | 22.2 | 66.7 | 11.1 | 0.473 |
| Providers' prerequisites | 16.7 | 20.8 | 62.5 | 35.3 | 0 | 64.7 | 27.8 | 11.1 | 61.1 | 58.3 | 16.7 | 25 | 66.7 | 0 | 33.3 | 0.047* |
| Evaluators' prerequisites | 12.5 | 8.3 | 79.2 | 35.3 | 17.6 | 47.1 | 44.4 | 5.6 | 50 | 50 | 33.3 | 16.7 | 55.6 | 0 | 44.4 | 0.016* |
| Recipients' prerequisites | 25 | 20.8 | 54.2 | 47.1 | 11.8 | 41.1 | 83.3 | 5.6 | 11.1 | 75 | 16.7 | 8.3 | 88.9 | 0 | 11.1 | 0.004* |
| Context's prerequisites | 79.2 | 0 | 20.8 | 70.6 | 5.9 | 23.5 | 77.8 | 11.1 | 11.1 | 58.3 | 16.7 | 25 | 44.4 | 11.1 | 44.4 | 0.412 |
| Global prerequisites of the whole intervention or for BeA | 4.2 | 83.3 | 12.5 | 11.8 | 76.4 | 11.8 | 22.2 | 77.8 | 0 | 25 | 75 | 0 | 11.1 | 77.8 | 11.1 | 0.488 |
| **Implementation characteristics of recipients' behaviors or actions (BeA)** |  |  |  |  |  |  |  |  |  |  |  |  |  |  |  |  |
| Global prerequisites for particular BeA | 4.2 | 83.3 | 12.5 | 29.4 | 58.8 | 11.8 | 38.9 | 61.1 | 0 | 58.3 | 41.7 | 0 | 66.7 | 33.3 | 0 | 0.009 |
| Providers' prerequisites | 20.8 | 4.2 | 75 | 35.3 | 5.9 | 58.8 | 38.9 | 0 | 61.1 | 75 | 0 | 25 | 66.7 | 11.1 | 22.2 | 0.049 |
| Evaluators' prerequisites | 12.5 | 0 | 87.5 | 41.2 | 0 | 58.8 | 50 | 0 | 50 | 75 | 0 | 25 | 77.8 | 0 | 22.2 | <.001* |
| Recipients' prerequisites | 25 | 4.2 | 70.8 | 47.1 | 0 | 52.9 | 83.3 | 5.6 | 11.1 | 91.7 | 0 | 8.3 | 100 | 0 | 0 | <.001* |
| Context prerequisites | 75 | 4.2 | 20.8 | 88.2 | 0 | 11.8 | 88.9 | 5.6 | 5.5 | 75 | 8.3 | 16.7 | 88.9 | 11.1 | 0 | 0.691 |
| Global characteristics of the implementation | 12.5 | 87.5 | 0 | 17.6 | 82.4 | 0 | 11.1 | 88.9 | 0 | 16.7 | 83.3 | 0 | 11.1 | 88.9 | 0 | 0.974 |
| Type of administration (e.g., remote, face-to-face) | 79.2 | 8.3 | 12.5 | 88.2 | 11.8 | 0 | 55.6 | 38.8 | 5.6 | 66.7 | 33.3 | 0 | 77.8 | 22.2 | 0 | 0.144 |
| Form of administration (e.g., individual, group) | 83.3 | 12.5 | 4.2 | 88.2 | 5.9 | 5.9 | 72.2 | 22.2 | 5.6 | 75 | 25 | 0 | 77.8 | 22.2 | 0 | 0.856 |
| Supervision/support (e.g., directed, autonomous, mixed) | 79.1 | 16.7 | 4.2 | 88.2 | 5.9 | 5.9 | 55.6 | 44.4 | 0 | 75 | 25 | 0 | 55.6 | 22.2 | 22.2 | 0.058 |
| Materials and support tools (e.g., multimedia, text, app) | 50 | 29.2 | 20.8 | 47.1 | 23.5 | 29.4 | 38.9 | 50 | 11.1 | 66.7 | 25 | 8.3 | 33.3 | 66.7 | 0 | 0.219 |
| Content and development of the sessions | 45.8 | 37.5 | 16.7 | 41.1 | 47.1 | 11.8 | 27.8 | 72.2 | 0 | 33.3 | 58.4 | 8.3 | 22.2 | 77.8 | 0 | 0.323 |
| Periodicity (time unit; e.g., once, daily, weekly, monthly) | 66.7 | 4.1 | 29.2 | 70.6 | 17.6 | 11.8 | 83.3 | 5.6 | 11.1 | 91.7 | 8.3 | 0 | 66.7 | 33.3 | 0 | 0.073 |
| Frequency per time unit (e.g., once, a number of times) | 70.8 | 4.2 | 25 | 64.7 | 17.6 | 17.7 | 72.2 | 16.7 | 11.1 | 83.3 | 8.3 | 8.4 | 55.6 | 33.3 | 11.1 | 0.497 |
| Total frequency (e.g., specific number, tailored) | 70.8 | 4.2 | 25 | 70.6 | 17.6 | 11.8 | 88.9 | 11.1 | 0 | 83.3 | 8.3 | 8.4 | 66.7 | 33.3 | 0 | 0.115 |
| Duration of sessions, dose or intensity (specific, tailored) | 62.5 | 4.2 | 33.3 | 58.8 | 35.3 | 5.9 | 50 | 27.8 | 22.2 | 58.4 | 8.3 | 33.3 | 44.5 | 33.3 | 22.2 | 0.180 |
| Geographical, spatial, physical and/or digital place | 91.7 | 4.1 | 4.2 | 82.4 | 17.6 | 0 | 83.3 | 16.7 | 0 | 91.7 | 8.3 | 0 | 100 | 0 | 0 | 0.627 |
| Moment in time (e.g., specific, tailored, irrelevant) | 91.7 | 0 | 8.3 | 100 | 0 | 0 | 100 | 0 | 0 | 83.3 | 0 | 16.7 | 100 | 0 | 0 | 0.177 |
| Social, cultural, political and/or economic setting | 91.7 | 4.1 | 4.2 | 94.1 | 5.9 | 0 | 94.4 | 5.6 | 0 | 75 | 8.3 | 16.7 | 100 | 0 | 0 | 0.410 |

As the number of BeA increased, the completeness of reporting improved. For example, in interventions with 2 BeA, reporting of global prerequisites increased to 11.8%, providers’ to 35.3%, and evaluators’ to 35.3%. By the time interventions addressed 3–4 BeA, global prerequisites were well described in 22.2% of studies, with evaluators’ (44.4%) and especially recipients’ prerequisites (83.3%) reported more often. This trend culminated in interventions with 4 BeA, where reporting of prerequisites was strongest: 25% for global prerequisites, 58.3% for providers, 50% for evaluators, and 75% for recipients. Context prerequisites remained relatively stable across groups (58–88%).

At the level of BeA (Fig. 10), the same pattern of improvement with increasing complexity was observed. For interventions with only one BeA, global prerequisites were reported in a replicable manner in 4.2% of cases, while providers’, evaluators’, and recipients’ prerequisites were accurately reported in 20.8%, 12.5%, and 25%, respectively. Again, context prerequisites were relatively well covered even in the simplest interventions (75%).

**Figure 10.**
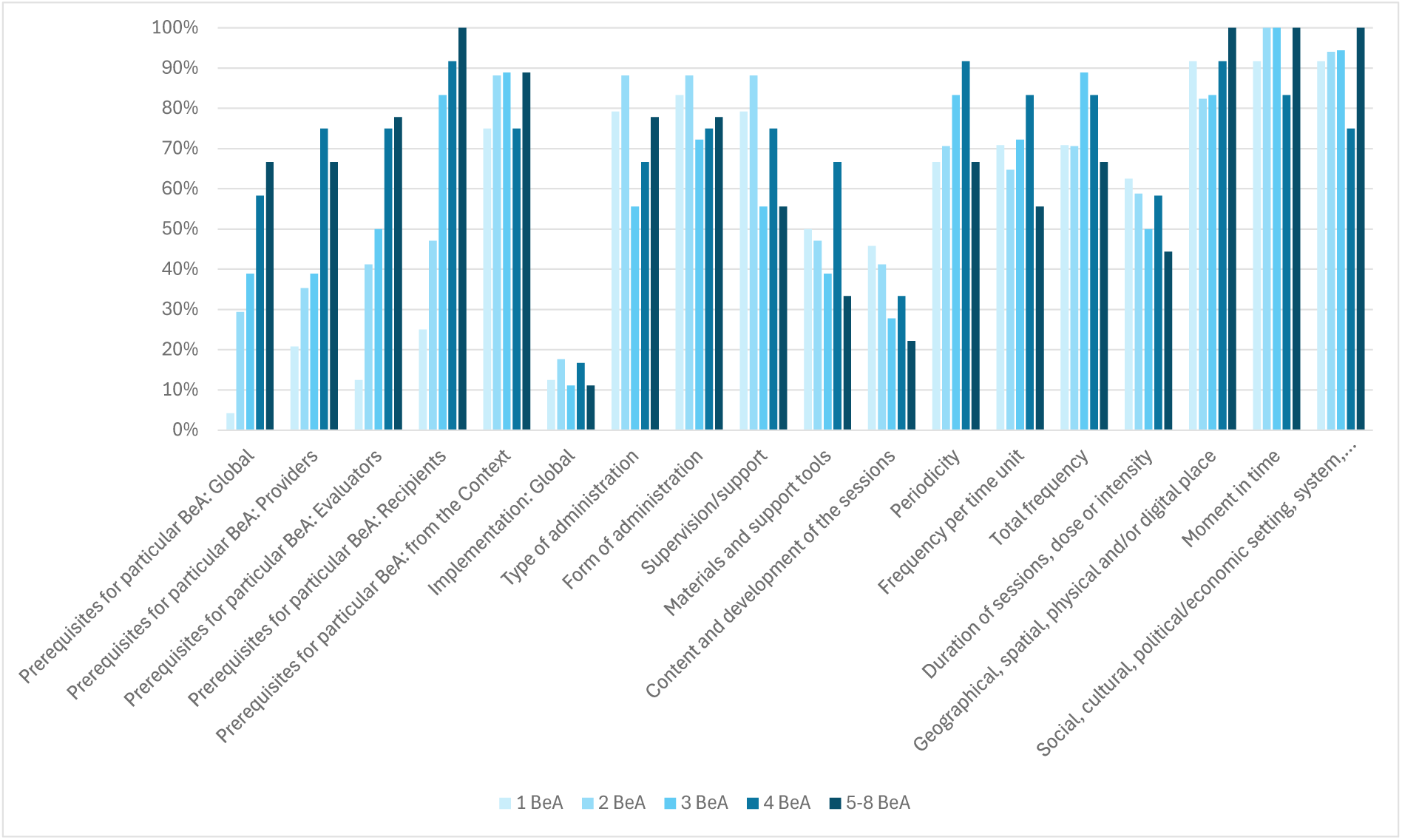
Percentage of implementation characteristics of the intervention’s recipients’ behaviors or actions (BeA) reported and/or replicable by number of BeA.

As the number of BeA increased, reporting became progressively more complete. With 2 BeA, global prerequisites well reported reached 29.4%, providers’ 35.3%, evaluators’ 41.2%, and recipients’ 47.1%, while context prerequisites climbed to 88.2%. For 3–4 BeA, adequate reporting was higher: 38.9% for global prerequisites, 50% for evaluators, and 83.3% for recipients. In interventions with 4 BeA, the reporting of prerequisites reached its peak, with 58.3% replicable for global prerequisites, 75% for providers and evaluators, 91.7% for recipients, and 75% for context. For the most complex interventions (5–8 BeA), reporting levels were also high, with recipients’ prerequisites accurately reported universally (100%) and evaluators’ prerequisites in 77.8%. Global implementation characteristics followed a slightly different pattern, generally showing low replicability (11.1–17.6%)

## Discussion

This scoping review mapped how psychological intervention reports describe implementation at both the whole-intervention level and at the level of recipients’ BeA across clinical, public health, social/organizational and educational fields. Only a minority of studies reported development-only objectives, while most combined development with evaluations of effectiveness and, to a lesser extent, feasibility. Few studies used a formal reporting quality tool or reported facilitators/barriers. Interventions typically targeted an individual-level single-category, with group and system level targeting being uncommon, and outcomes were generally well specified. Reporting of the implementation sequence was generally adequate, yet structured representations such as tables, lists or diagrams were underused, and costs were rarely reported. At the BeA level, generic structural features (such as format, supervision, periodicity, setting and timing) were usually described in a replicable way, whereas content and materials, adaptation or customization, and especially fidelity or adherence were often incompletely reported. Relevant aspects of complexity such as multi-level recipients, moderation or mediation effects were seldom analysed, although reporting quality tended to improve as the number of BeA increased.

Our findings show that, despite decades of general reporting guidelines (CONSORT, STROBE, PRISMA, SPIRIT, TREND) and intervention-specific reporting tools (TIDieR, CReDECI2, GUIDED, iCAT_SR, RIPIf), their adoption remains limited and key implementation details continue to be omitted. This is consistent with previous works showing that endorsement of reporting tools and guidelines alone does not guarantee adherence and complete reporting (Turner et al., 2012; Bastuji-Garin et al., 2013; Clayson et al., 2019). Similarly, key recommendations from the MRC guidance on developing and evaluating (complex) interventions have been largely overlooked, as most studies report both intervention development and testing within a single paper, which tends to compromise the replicability of the intervention.

Our results also resonate strongly with TIDieR’s emphasis on specifying *what* was delivered (materials/content) and *how* (dose, mode, procedures). The comparatively weak reporting for content/materials (and moderate reporting for dose/intensity) indicates that, even when structural aspects are reported, some core elements of interventions are often left implicit.

Contemporary frameworks and tools such as MRC and iCAT_SR ask authors to articulate how components are expected to work (mechanisms), for whom, in what contexts, and with what interdependencies. The rarity of multi-level targeting and explicit tests of mediation/moderation among BeA suggests that many reports still treat interventions largely as packages rather than as systems of interacting parts. Explicitly modelling and reporting inter-component relations, and using logic models to link BeA to outcomes, would align practice with these frameworks and support richer cumulative science (Moore et al., 2015).

Two additional gaps stand out. First, implementation fidelity was rarely reported in a replicable way. The conceptual framework of Carroll et al. (2007) describes fidelity as a multidimensional construct (adherence, dose, quality of delivery, participant responsiveness, differentiation) that should be planned, measured, and reported systematically; the pattern we observed suggests these dimensions remain underspecified in psychological intervention reports. Moreover, meta-research consistently shows that implementation quality is tied to effect sizes, reinforcing the importance of robust fidelity reporting (Durlak & DuPre, 2008). Second, costs were almost never reported, yet budget impact and cost-effectiveness information are essential to scaleup and policy translation. The CHEERS statement (Husereau et al., 2022) provides explicit, modernized guidance on transparent reporting of economic evaluations and is broadly applicable across intervention types; the near absence of cost reporting in our sample signals an actionable opportunity for improvement in future reports. From a practice standpoint, insufficient reporting of fidelity, adaptation policies, and costs undermines implementation planning and weakens the claim that a delivered service is truly evidence based. The taxonomy of Proctor et al. (2011) clarifies that feasibility, fidelity, penetration, sustainability, and cost are implementation outcomes in their own right; adequate reporting of these domains is therefore necessary both for replication and for implementation science to cumulate knowledge.

Reporting quality is also greatly enhanced when the structure and contents of a scientific report follow widely accepted standards (e.g., APA). In this regard, some inconsistencies have also been found in this review. Critical prerequisites and contextual details are frequently reported outside the Methods section, impeding discoverability and machine readability for evidence synthesis. Consolidating these items in Methods (with stable supplementary files hosting manuals, checklists, and materials) would align with SPIRIT/CONSORT transparency goals and facilitate automated screening by meta-research tools.

However, two encouraging findings emerge from our study. First, the implementation sequence was often reported with sufficient detail, and in more complex interventions, the reporting of prerequisites (especially for recipients and evaluators) improved. Second, generic structural elements (e.g., type/form of administration, supervision, settings) and timing-related characteristics (e.g., periodicity, frequency, duration) were frequently replicable. These strengths provide a solid foundation to build upon.

### Limitations and strengths

This scoping review aimed to map reporting practices rather than quantify causal relationships or effectiveness. The review timeframe encompasses all major recent reporting frameworks, guidelines, and tools. Although selection was randomized within fields, the sample size per field limits statistical power to detect small differences and may not fully capture sub-disciplinary variations. Judgments of “replicable” versus “reported but not replicable” required coder interpretation; we mitigated this through piloting, independent double extraction, and consensus procedures, achieving a substantial interrater agreement at full-text screening. Finally, the theoretically guided and empirically grounded data extraction instrument developed for this study represents a key output. It provides powerful insights into the conceptualization of interventions’ active ingredients (BeA), and a comprehensive list of appraisable properties of both these BeA and the whole intervention.

### Conclusions and future directions

Reporting of psychological interventions shows encouraging coverage of structural “when/where/how” elements and implementation sequences, but persistent weaknesses in what (content/materials), how well (fidelity), whether and how it changes (adaptation/customization), and how much it costs (economic data). These deficiencies blunt replication, weaken cumulative syntheses, and impede implementation at scale, precisely the problems the reporting guideline ecosystem was designed to solve. Increasing the promotion of intervention reporting tools and guidelines, such as TIDieR, CONSORT-SPI, RIPI-f, GUIDED, and CReDECI-2, in psychological intervention reports, alongside intervention development and evaluation frameworks and tools (e.g., MRC and iCAT_SR), and economic reporting standards (e.g., CHEERS 2022), would help address the most consequential gaps identified in our scoping review. Additionally, using content taxonomies, such as the Behavior Change Technique Taxonomy (BCTTv1; (Michie et al., 2013)), can sharpen the granularity and comparability of content reporting via a more precise specification of active intervention components and thereby strengthen replication and synthesis.

Our results suggest that future research should focus primarily on:

a. Editorial interventions: evaluate editorial interventions, such as mandatory checklists, structured tables or diagrams, and automated report audits, which have demonstrated improvements in research integrity in other domains and adapting them to the level of granularity required for BeA.
b. Level of BeA: jointly develop a minimum set of BeA characteristics, linking each BeA to prerequisites, materials or content, procedures, dosage, fidelity measures, and outcomes, including hypothesized mediation and moderation effects between BeA.
c. Complexity: increase the routine use of logic models and process evaluations to assess the performance of components across different contexts, including multilevel recipients and intercomponent pathways.
d. Fidelity measurement and reporting: prospectively adopt multicomponent frameworks and evaluate whether improved fidelity reporting enhances effect estimates and reduces heterogeneity in meta-analyses.
e. Economic transparency: integrate CHEERS 2022 elements, such as resource use, costs, and analytical perspective, into intervention reports, even when a full economic evaluation is not conducted, to better inform scaling-up decisions.

Beyond the scope of our findings, future research should explore the potential role of artificial intelligence (AI) in assessing reporting quality in intervention studies. Domain-specific language models such as BioBERT and PubMedBERT as well as recent reporting-focused work (e.g., Chen et al., 2025) suggest promising avenues for an AI-assisted appraisal of reporting quality.

## Data availability statement

The data supporting the findings of this study are openly available in the Open Science Framework repository at https://doi.org/10.34810/data2792.

## Funding

This work was supported by Grant PID2022-141403NB-I00 funded by the Spanish Government (MCIN/ AEI/10.13039/501100011033/FEDER, UE), and 2021SGR-00806, funded by the Government of Catalonia (Spain). The authors declare no conflict of interest, and the funders had no role in the study design, data collection, analysis, decision to publish, or preparation of the manuscript. No additional external funding was received for this study.

## Competing interests

The authors have declared that no competing interests exist.

## Acknowledgements

An AI language model (Perplexity AI) was used to assist in correcting typographical and grammatical errors and to improve the clarity and fluency of English writing. The authors reviewed and approved all content.

## APPENDIX A. Search strategy

### Public health

*PsycInfo (2609)*

((((title((“intervention*” OR “treatment*” OR “therap*” OR “program*”) NEAR/5 (“develop*” OR “protocol” OR “feasibility” OR “effica*” OR “effectiv*”)) NOT title(“review*” OR “meta-anal*”)) AND PEER(yes))) AND pd(20140101-20241231)) AND ccl.exact(“Health & Mental Health Treatment & Prevention” OR “Promotion & Maintenance of Health & Wellness” OR “Health & Mental Health Services” OR “Health Psychology & Medicine” OR “Home Care & Hospice” OR “Sports & Exercise” OR “Childrearing & Child Care” OR “Nursing Homes & Residential Care” OR “Health & Mental Health Personnel Issues” OR “Aging & Older Adult Development”)

*Web of Science (677)*

((((TI=((“intervention*” OR treatment* OR therap* OR program*) NEAR/5 (develop* OR protocol OR feasibility OR effica* OR effectiv*))) NOT TI=(review* OR meta-anal*)) AND DOP=(2014-01-01/2024-12-31)) AND DT=(Article)) AND (TASCA==(“PSYCHIATRY” OR “PSYCHOLOGY”) AND TASCA==(“SUBSTANCE ABUSE” OR “GERIATRICS GERONTOLOGY” OR “NURSING” OR “PEDIATRICS” OR “GERONTOLOGY” OR “HEALTH CARE SCIENCES SERVICES” OR “PUBLIC ENVIRONMENTAL OCCUPATIONAL HEALTH” OR “HEALTH POLICY SERVICES” OR “NUTRITION DIETETICS” OR “FAMILY STUDIES” OR “ONCOLOGY” OR “REHABILITATION” OR “HOSPITALITY LEISURE SPORT TOURISM” OR “SPORT SCIENCES” OR “SURGERY” OR “IMMUNOLOGY” OR “OBSTETRICS GYNECOLOGY”))

### Clinical

*PsycInfo (3242)*

((((title((“intervention*” OR “treatment*” OR “therap*” OR “program*”) NEAR/5 (“develop*” OR “protocol” OR “feasibility” OR “effica*” OR “effectiv*”)) NOT title(“review*” OR “meta-anal*”)) AND PEER(yes))) AND pd(20140101-20241231)) AND ccl.exact(“Cognitive Therapy” OR “Psychotherapy & Psychotherapeutic Counseling” OR “Specialized Interventions” OR “Behavior Therapy & Behavior Modification” OR “Affective Disorders” OR “Neurodevelopmental & Autism Spectrum Disorders” OR “Anxiety Disorders” OR “Behavior Disorders & Antisocial Behavior” OR “Behavioral & Psychological Treatment of Physical Illness” OR “Psychological & Physical Disorders” OR “Schizophrenia & Psychotic States” OR “Psychological Disorders” OR “Eating Disorders” OR “Speech & Language Therapy” OR “Interpersonal & Client Centered & Humanistic Therapy” OR “Personality Disorders” OR “Psychoanalytic Therapy”)

*Web of Science (1788)*

((((TI=((“intervention*” OR treatment* OR therap* OR program*) NEAR/5 (develop* OR protocol OR feasibility OR effica* OR effectiv*))) NOT TI=(review* OR meta-anal*)) AND DOP=(2014-01-01/2024-12-31)) AND DT=(Article)) AND (TASCA==(“PSYCHOLOGY CLINICAL”) OR (TASCA==(“PSYCHIATRY” OR “PSYCHOLOGY”) AND TASCA==(“CLINICAL NEUROLOGY” OR “PHARMACOLOGY PHARMACY” OR “PSYCHOLOGY PSYCHOANALYSIS” OR “GENETICS HEREDITY”)))

### Social and organizational

*PsycInfo (865)*

((((title((“intervention*” OR “treatment*” OR “therap*” OR “program*”) NEAR/5 (“develop*” OR “protocol” OR “feasibility” OR “effica*” OR “effectiv*”)) NOT title(“review*” OR “meta-anal*”)) AND PEER(yes))) AND pd(20140101-20241231)) AND ccl.exact(“Community & Social Services” OR “Professional Personnel Attitudes & Characteristics” OR “Behavior Disorders & Antisocial Behavior” OR “Occupational & Vocational Rehabilitation” OR “Social Processes & Social Issues” OR “Organizational Psychology & Human Resources” OR “Criminal Behavior & Juvenile Delinquency” OR “Personnel Management & Selection & Training” OR “Working Conditions & Industrial Safety” OR “Management & Management Training” OR “Organizational Behavior” OR “Personnel Attitudes & Job Satisfaction” OR “Social Psychology” OR “Mass Media Communications” OR “Marriage & Family” OR “Occupational Interests & Guidance” OR “Police & Legal Personnel” OR “Consumer Attitudes & Behavior”)

*Web of Science (1098)*

((((TI=((“intervention*” OR treatment* OR therap* OR program*) NEAR/5 (develop* OR protocol OR feasibility OR effica* OR effectiv*))) NOT TI=(review* OR meta-anal*)) AND DOP=(2014-01-01/2024-12-31)) AND DT=(Article)) AND (TASCA==(“SOCIAL SCIENCES INTERDISCIPLINARY” OR “SOCIAL WORK” OR “SOCIAL SCIENCES BIOMEDICAL” OR “SOCIAL ISSUES” OR “SOCIAL SCIENCES MATHEMATICAL METHODS” OR “MANAGEMENT” OR “CRIMINOLOGY PENOLOGY”))

### Education

*PsycINFO (1489)*

((((title((“intervention*” OR “treatment*” OR “therap*” OR “program*”) NEAR/5 (“develop*” OR “protocol” OR “feasibility” OR “effica*” OR “effectiv*”)) NOT title(“review*” OR “meta-anal*”)) AND PEER(yes))) AND pd(20140101-20241231)) AND ccl.exact(“Curriculum & Programs & Teaching Methods” OR “Professional Education & Training” OR “Educational Administration & Personnel” OR “Educational/Vocational Counseling & Student Services” OR “Educational & School Psychology” OR “Special & Compensatory Education” OR “Classroom Dynamics & Student Adjustment & Attitudes” OR “Developmental Psychology”)

*Web of Science (589)*

((((TI=((“intervention*” OR treatment* OR therap* OR program*) NEAR/5 (develop* OR protocol OR feasibility OR effica* OR effectiv*))) NOT TI=(review* OR meta-anal*)) AND DOP=(2014-01-01/2024-12-31)) AND DT=(Article)) AND (TASCA==(“PSYCHOLOGY DEVELOPMENTAL”) OR (TASCA==(“PSYCHIATRY” OR “PSYCHOLOGY”) AND TASCA==(“EDUCATION SPECIAL” OR “EDUCATION EDUCATIONAL RESEARCH”)))

## Notes

### Competing Interest Statement

The authors have declared no competing interest.

### Summary of Updates

The search strategy details for the method were updated. The formatting of Figures 1a, 1b, and 2, and Table 1 was updated. An appendix with the search syntax was added.

